# Pancreatic enzymes and/or bile acids for acutely ill severely malnourished children – a double-blind randomized placebo-controlled trial

**DOI:** 10.64898/2025.12.22.25342418

**Authors:** Robert H. J. Bandsma, Mohammod Jobayer Chisti, Wieger P. Voskuijl, Moses M. Ngari, Ezekiel Mupere, Isaiah Njagi, Johnstone Thitiri, Isabel Potani, Emmie Mbale, Dennis Chasweka, Chikondi Makwinja, Chisomo Eneya, Celine Bourdon, Abu Sadat Mohammad Sayeem Bin Shahid, Shamsun Nahar Shaima, Gazi Md Salahuddin Mamun, Christina L. Lancioni, Christopher Maronga, Christopher Lwanga, Michael Atuhairwe, Kirkby D. Tickell, Caroline Tigoi, Moses Mburu, Narshion Ngao, Alex Dmitrienko, Farzana Afroze, Lubaba Shahrin, Tahmeed Ahmed, Judd L. Walson, James A. Berkley

## Abstract

**Background:** Translocation of bacteria across the intestinal barrier has been postulated to contribute to mortality among severely malnourished children. Pancreatic enzymes (PE) and bile acids (BA) have anti-bacterial properties in the small intestine, but severe malnutrition is associated with impaired exocrine pancreatic and hepatobiliary functions. We evaluated whether ancillary treatment with PE and BA improves survival in hospitalized, acutely ill, severely malnourished children.

**Methods and findings:** We conducted a multicenter, adaptive, 2×2 factorial, randomized, double-blinded controlled trial of PE and BA (ursodeoxycholic acid) administered for 21 days in hospitalized children with severe malnutrition. The primary outcome was mortality within 60 days of enrollment. Secondary outcomes included the incidence of Serious Adverse Events (SAEs); grade 3 or 4 toxicity events before day 21; number of days with diarrhoea; use of second- or third-line antibiotics during the admission; duration of initial hospitalization; and changes in anthropometry from enrolment to follow-up at 21 and 60 days after start of the intervention.

We enrolled 429 children (median age 10.4 months (interquartile range (IQR) 4.8 18.0)). Participants were randomized to PE (n=213, 50%) or PE-placebo (n=216, 50%) and to BA (n=212, 49%) or BA-placebo (n=217, 51%). Mortality by day 60 did not differ between children assigned to PE compared to PE-placebo (36 children (17%) vs 35 (16%)), irrespective of being assigned BA or BA-placebo (Hazard ratio (HR) 1.02 (95% CI 0.64 1.62), P=0.94). Also, 39 children (18%) assigned to BA died compared to 32 (15%) with BA-placebo, irrespective of PE/PE-placebo (HR 1.27 (95%CI 0.79 2.02), P=0.32). The trial was stopped early based on futility. There were no significant differences in secondary outcomes, other than children assigned to BA spent fewer days on second-line antibiotics than those receiving BA-placebo.

**Conclusions:** Pancreatic enzymes and/or ursodeoxycholic acid do not reduce mortality, serious adverse events, or improve other clinical outcomes in hospitalized, acutely ill, severely malnourished children.

**Funding:** The Bill and Melinda Gates Foundation. Pancreatic enzymes were provided at subsidized cost by Abbott Laboratories, Illinois, Untied States and ursodeoxycholic acid was provided in kind by Opsonin Pharma Ltd., Dhaka, Bangladesh.

## Introduction

The World Health Organization (WHO) provides standardized protocols for the inpatient management of severely malnourished children with acute illness or complications, including specialized feeding and antibiotic therapy. However, mortality rates remain unacceptably high, prompting a pressing search for new evidence-based interventions ^1,2^. Despite this urgency, scientific understanding of the pathophysiology driving mortality remains limited. Robust clinical trials, crucial for advancing the management of severely ill malnourished children, are lacking.

Maintaining a balanced and compartmentalized intestinal microbiota is essential for overall health. However, this balance is disrupted in children with severe malnutrition resulting in significant dysbiosis, including colonization of the upper small intestine (overgrowth) with pathogenic Enterobacteriaceae ^3–5^. Dysbiosis and small intestinal bacterial overgrowth (SIBO) are associated with intestinal inflammation, compromised barrier function and impaired growth ^6,7^. Moreover, intestinal dysfunction has been associated with increased mortality in severely malnourished children, potentially due to bacterial translocation from the upper small intestinal lumen into the bloodstream ^8^. Although empiric antibiotics are strongly recommended for hospitalized children with severe acute malnutrition, mortality linked to infections remain high. While metronidazole or amoxicillin-clavulanic acid are used in management of SIBO and dysbiosis, evidence supporting their use in severely malnourished children is limited. Also, growing concerns around antibiotic resistance (AMR) and further disruption of the microbiome highlight the need for adjunctive therapies that target the underlying pathophysiology to improve treatment efficacy.

The exocrine pancreas and the liver are essential for nutrient digestion and absorption and their secretions also directly regulate the intestinal microbiota. Pancreatic acinar cells secrete enzymes such as trypsin, lipase and amylase that break down proteins, lipids and carbohydrates. Meanwhile, hepatic cholangiocytes produce bile containing bile acids, which form micelles that aid lipases, thus, promoting effective absorption. Beyond their digestive functions, pancreatic enzymes and bile acids regulate bacteria in the upper small intestine through several mechanisms including the action of antibacterial peptides, phospholipids and detergent-like properties, therefore playing a critical role in regulating SIBO and dysbiosis ^9–15^. A recent study using microbiome-directed complementary foods (MDCF) has suggested that addressing dysbiosis directly in malnourished children can be beneficial and promote growth ^16^. Severely malnourished children have exocrine pancreatic insufficiency and hepatobiliary dysfunction that alters bile acid metabolism ^17–19^. A recent pilot trial providing pancreatic enzyme supplementation to inpatient children with complicated severe malnutrition suggested a potential reduction in mortality ^20^. No trials of bile acid supplementation have been undertaken in this population, although pre-clinical studies have indicated potential beneficials ^21,22^.

We hypothesized that providing pancreatic enzymes and ursodeoxycholic acid could reduce mortality in acutely ill severely malnourished children. We therefore conducted a randomized clinical trial to test the effects of exocrine pancreatic enzyme and/or bile acid supplementation on mortality in these children.

## Methods

### Study design and setting

We conducted a multicenter, adaptive clinical trial using a 2×2 factorial, randomized, double-blind, placebo-controlled design to evaluate two interventions. This design was chosen to evaluate the individual and combined effects of the interventions, facilitating their assessment as a potential integrated ‘care package’. The clinical trial was designed and conducted under the umbrella of the Childhood Acute Illness and Nutrition (CHAIN) Network (https://chainnetwork.org/). There was no direct patient or public involvement in the design of this trial. The study took place in five research sites in four countries (Kenya, Uganda, Bangladesh and Malawi) and received ethical approval from the following bodies: 1) the KEMRI Scientific and Ethical Review Unit (SERU), Nairobi, Kenya; 2) School of Biomedical Sciences, the Research and Ethics Committee Makerere University College of Health Sciences and the Uganda National Council for Science and Technology (UNCST), Uganda; 3) the Oxford Tropical Research Ethics Committee (OxTREC), University of Oxford, UK; 4) College of Medicine Research and Ethics Committee (COMREC), Kamuzu University of Health Sciences, Malawi; 5) Hospital for Sick Children, Toronto, Canada and 6) International Centre for Diarrheal Disease Research (icddr,b), Bangladesh. The trial was registered on an independent study registry (www.clinicaltrials.gov, NCT04542473) and all case report forms and standard operating procedures are publicly available at https://chain.tghn.org/chain_pb_sam/. A parent or legal guardian provided written informed consent before each child’s enrollment. The trial adhered to the International Council for Harmonisation of Technical Requirements for Pharmaceuticals for Human Use (ICH), Guideline for Good Clinical Practice (GCP) and the Declaration of Helsinki. Oversight was provided by an independent Trial Steering Committee and Data Safety Monitoring Board. Reporting was conducted according to the most recent CONSORT statement. No significant changes to the protocol were made after initiation of the trial.

### Participants

Children were recruited from the following sites: Kilifi County Hospital, Kenya; Coast General Hospital, Mombasa, Kenya; Mbagathi Sub-County Hospital, Nairobi, Kenya; Queen Elizabeth Central Hospital (QECH) Blantyre, Malawi; Dhaka Hospital of the icddr,, Dhaka, Bangladesh; and Mulago National Referral Hospital, Kampala, Uganda.

Screening, recruitment, consent, and enrollment took place during the hospital admission process or within 72 hours of admission. Screening was based on WHO criteria for severe malnutrition (weight-for-length/height z-score (WHZ) <-3 and/or mid upper arm circumference (MUAC) <115mm (or <110 mm for children aged below 6 months), or presence of nutritional, bilateral pitting oedema)^1^. Additional trial-specific criteria were then applied*. Inclusion criteria:* age >2 to <59 months; admitted to hospital with an acute, non-traumatic illness and enrolled within 72 hours of admission; presence of two or more features of childhood illness-danger signs as specified in **Supplementary Table 1**; accompanied by a caregiver that provided written informed consent; primary caregiver planning to stay within the study area for the study duration. *Exclusion criteria:* requires immediate cardiac/respiratory resuscitation; terminal illness (other than severe acute malnutrition) likely to result in death within 6 months, as judged by the recruiting clinician; known congenital heart disease; admission for trauma or surgery; known liver or exocrine pancreatic disorders – e.g. biliary atresia, history of gallstones, cystic fibrosis or clinical jaundice, known stomach or duodenal ulcer; known intolerance or allergy to any study medication; residence outside the hospital’s catchment area.

### Study interventions

In addition to standard in-hospital care, severely malnourished children were randomised in a 2×2 factorial design to receive study treatments. *Randomization 1:* oral/enteral pancreatic enzymes: lipase, amylase and protease (PE) or matching placebo (PE-placebo). *Randomization 2:* oral/enteral bile acids: ursodeoxycholic acid (BA) or matching placebo (BA-placebo). The pancreatic enzymes (Creon^®^ gastro-resistant pellets, Abbott Laboratories, Neustadt, Germany) and PE-placebo (Abbott Laboratories) were provided as a visually similar granulate. The ursodeoxycholic acid (Liconor^®^ suspension, Opsonin Pharma Limited, Barishal, Bangladesh) and BA-placebo (Opsonin Pharma Limited) was provided as matching formulation, identically packaged in dark-colored bottles. Children received i) PE or PE-placebo and ii) BA or BA-placebo for a total duration of 21 days. For children not able to feed orally while hospitalized, treatments were administered enterally via a nasogastric tube. The target dose of pancreatic enzymes was 3000 IU lipase/kg, twice per day, just before a feed. This dose (1440 IU/kg amylase and 80 IU/kg protease) is recommended for children with cystic fibrosis or exocrine pancreas insufficiency ^23^. Given the sachet-based formulation, dosing followed internationally accepted weight bands, allowing a range of 2000 to 4000 IU/kg/day. Ursodeoxycholic acid was administered at a dose of 10 mg/kg twice per day before feeding using a 50 mg/ml suspension^24^. For children with nutritional oedema, the body weight used for dosing was pragmatically reduced by 10%. Children assigned to PE- and/or BA-placebo arms received equivalent volumes to counterpart formulations.

### Data collection

Demographic, clinical and pre-enrolment care data were recorded at enrolment by study clinicians. Daily assessments were performed throughout hospitalization, and at discharge, and during two follow-up visits at days 21 and 60 after enrolment. Caregivers were encouraged to contact the study team and return to the study hospital if the child deteriorated or if parents had concerns about their child’s health. To investigate reasons for readmission, participants who were re-admitted to hospital underwent standardised clinical assessment and additional investigations as per hospital and national guidelines. For those re-admitted to non-study hospitals, the date of readmission and discharge, and clinical diagnosis were recorded. Re-admission was considered a serious adverse event (SAE). Post-discharge interventional product adherence was assessed indirectly by comparing the weight of each medication bottle dispensed to caregivers at discharge with the weights of the same bottle upon return at the day 21 visit. The difference in weight was considered to represent the amount of product consumed, which was compared against the prescribed target dose to estimate adherence. These calculations were restricted to children who were discharged prior to day 21 and survived to attend that follow-up visit.

### Outcomes

The primary endpoint for both interventions was mortality within 60 days of enrolment. Secondary outcomes were:

- Serious Adverse Events of any cause within 60 days
- Toxicity: grade 3 or 4 toxicity events from enrolment to day 21 (i.e., end of the intervention)
- Number of days with diarrhoea
- Number of days with clinical features of sepsis (defined as any WHO danger sign, shock, weak pulse, capillary refill time >3 seconds or temperature gradient)
- Use of second- and third-line antibiotics during index admission and any readmission
- Number of days from enrolment (index admission) to discharge
- Change in MUAC, weight-for-length (WLZ), weight-for-age (WAZ) and length-for-age (LAZ) from enrolment to 21 and 60 days

### Randomisation and masking

Randomisation was stratified by age group, site and pre-assigned in randomly sized blocks per site. The randomisation sequence was generated by an independent statistician at the KEMRI Wellcome Trust Research Programme, ensuring allocation concealment. The randomization list was sent to the manufacturer who prelabeled the products. The products were shipped to the sites with the participant number.. Eligible participants were assigned to treatment arms using the participant number. The trial was double-blinded. Investigational products and placebo formulations for both interventions were matched in appearance, volume and packaging. Randomization codes linking study numbers to treatment allocation were securely held by an off-site trial statistician and were not accessible until the database was unlocked. Unblinding was permissible only in the event of a medical emergency or serious safety concern where knowledge of allocation would inform patient care and deemed in the best interest of the child. In such cases, the site principal investigator could request unblinding through a defined procedure. Allocation remained concealed from participants, caregivers, and trial staff, including the trial statistician.

### Statistical methods

#### Sample size

The sample size was determined to detect a reduction in 60-day mortality from 24% to 16%, corresponding to a hazard ratio (HR) of 0.66. This estimate was based on findings from a pilot trial evaluating pancreatic enzyme supplementation in acutely ill children with severe malnutrition and mortality rates reported in this population by a recent large observational study conducted by our group ^2^. For a power of 90%, two-tailed alpha of 5%, and 5% loss to follow-up, a total sample size of 1200 was required (i.e., 300 children per treatment arm).

### Interim and final analyses

Two interim analyses were conducted, with decisions to continue or discontinue the trial based on prespecified criteria using Bayesian methods ^25^. A common prior distribution was assumed for the incidence rates across the trial arms and was derived as follows. A composite event defined as the occurrence of any SAEs, including death and readmission, or grade 3 or 4 toxicity was developed. The time to this composite event (defined as the occurrence of any SAEs, including death and readmission, or grade 3 or 4 toxicity) was assumed to follow a Weibull distribution with a shape parameter of 0.5 and scale parameters determined from historical data. The scale parameter for deaths was derived under the assumption that a 60-day mortality rate is 25% and the scale parameter for SAEs and non-fatal Grade 3 or 4 events was computed assuming a 60-day event rate of 12.5%. The mean of the common prior distribution was set to 0.19, the resulting incidence rate (expected number of composite events per subject per month). Using the derived mean incidence rate and an assumed coefficient of variation (0.3), the common prior distribution of the incidence rates was a gamma distribution with the shape parameter 11.1 and rate parameter 58.5. A clinically meaningful non-inferiority margin was set at 10% of the assumed incidence rate, i.e., 0.019 at Interim analysis 1. The first interim analysis was conducted after enrolling 200 children and assessed non-inferiority of each intervention compared to placebo using the composite endpoint. An intervention was retained if the posterior probability of non-inferiority exceeded 0.8 (80%). After enrollment of 429 participants, the second interim analysis assessed the superiority of each intervention compared to placebo using the same composite endpoint. An intervention was retained if the posterior probability of superiority reached or exceeded 80%. An independent Data Monitoring and Safety Board (DMSB) committee reviewed both interim analysis and provided recommendations to continue or stop the trial.

The final analyses were conducted on the intention-to-treat (ITT) population, including all randomized children. We used factorial analysis where each intervention was analysed separately and compared to their respective placebo groups for reasons of efficiency. To investigate interaction between the two interventions, we compared models with and without intervention interaction term using likelihood ratio test. For the primary outcome 60-day mortality, the absolute number of deaths, mortality rates and corresponding 95% confidence intervals (CI), are reported for each arm. Treatment efficacy was assessed using difference in mortality rates and HR of death within 60 days of enrolment. The HR were estimated using Cox proportional hazards regression, with confirmation of proportionality. Survival was visualized using Kaplan-Meier curves. A sensitivity analysis was performed with recruiting site as a random-effect to account for potential site-level variation. Causes of death, as adjudicated by an independent endpoint review panel, are reported by trial arms. As sensitivity analyses, the baseline characteristics and primary outcome are reported stratified by the four treatment allocations (Pancreatic Enzymes only, Bile acid only, Pancreatic Enzymes + Bile acid and double placebo).

For prespecified secondary outcomes, number of episodes and incidence rates of all SAEs were estimated and compared between intervention groups using incidence rate ratios (IRRs). Incidence rates of hospital readmissions were similarly analyzed. Episodes and incidence rates of suspected grade 3 and 4 toxicity event during the 21 days of intervention were also reported and compared using IRRs. Days with sepsis, diarrhoea, second- and third-line antibiotics, were reported and compared between interventions using zero-inflated negative binomial regression models. This approach accounted for over-dispersion, and the high frequency of zero counts. Days spent in hospital were reported as median with interquartile range (IQR) and compared between groups using negative binomial regression. Changes in anthropometric z-scores from baseline to 21 and 60 days were compared between groups using linear regression models, adjusted for absolute baseline values.

## Results

### Dates, sites and baseline characteristics

Between June 20, 2021, and October 17, 2022, a total of 429 children were enrolled. Of these, 213 (50%) were randomly assigned to receive PE and 216 (50%) received PE-placebo; while 212 (49%) were assigned to BA and 217 (51%) to BA-placebo. Their overall median age was 10.4 months (IQR 4.8 18.0) and 234 (55%) children were male. Site enrollment was as follows: 119 (28%) children in Blantyre Malawi, 117 (27%) in Dhaka Bangladesh, 84 (20%) in Kampala Uganda, 94 (22%) in Kilifi Kenya, and 15 (3.5%) in Nairobi Kenya. At enrolment, the cohort had a mean (SD) MUAC of 10.3 (1.19) cm; WHZ -3.44 (1.38) and HAZ -3.61 (1.75). Nutritional oedema was present in 103 (24%) children. At admission, the most common diagnoses were gastroenteritis (246, 57%), severe pneumonia (78, 18%), malaria (39, 9.1%) and HIV seropositivity (39, 9.1%). Baseline demographic and clinical characteristics were similar between randomized groups (**Table 1**). Baseline characteristics stratified by the four treatment allocations are shown in **S2 Table.**

**Table 1.** Baseline characteristics.

|  | Pancreatic enzymes randomization |  | Bile acid randomization |  |
| --- | --- | --- | --- | --- |
|  | Placebo (n=216) | Pancreatic enzymes (n=213) | Placebo (n=217) | Bile acid (n=212) |
| Age in months; median (IQR) | 10.6 (5.1–16.7) | 9.9 (4.6–18.9) | 10.2 (4.7–17.3) | 10.5 (5.2–20.4) |
| Sex |  |  |  |  |
| Female | 91 (42) | 104 (49) | 102 (47) | 93 (44) |
| Small birth size | 64 (30) | 66 (31) | 63 (29) | 67 (32) |
| Previous hospital admission | 78 (36) | 76 (36) | 72 (33) | 82 (39) |
| Reported antibiotic in 7 days prior to admission | 89 (41) | 89 (42) | 94 (43) | 84 (40) |
| Currently breastfeeding | 89 (41) | 92 (43) | 104 (48) | 77 (36) |
| Attending outpatient nutrition program prior to admission |  |  |  |  |
| No | 186 (86) | 184 (86) | 190 (88) | 180 (85) |
| Supplementary/Therapeutic | 30 (14) | 29 (14) | 27 (12) | 32 (15) |
| Site |  |  |  |  |
| Blantyre, Malawi | 59 (27) | 60 (28) | 60 (28) | 59 (28) |
| Dhaka, Bangladesh | 59 (27) | 58 (27) | 60 (28) | 57 (27) |
| Kampala, Uganda | 43 (20) | 41 (19) | 43 (20) | 41 (19) |
| Kilifi, Kenya | 46 (21) | 48 (23) | 47 (22) | 47 (22) |
| Nairobi, Kenya | 9 (4.2) | 6 (2.8) | 7 (3.2) | 8 (3.8) |
| Anthropometry: mean ±sd | | | | |
| MUAC | 10.3 ±1.15 | 10.3 ±1.24 | 10.2 ±1.01 | 10.4 ±1.35 |
| WHZ | -3.49 ±1.36 | -3.38 ±1.40 | -3.35 ±1.37 | -3.53 ±1.39 |
| WAZ | -4.56 ±1.15 | -4.63 ±1.08 | -4.49 ±1.06 | -4.71 ±1.17 |
| HAZ | -3.59 ±1.84 | -3.62 ±1.66 | -3.53 ±1.70 | -3.68 ±1.80 |
| Oedema | 54 (25) | 49 (23) | 43 (20) | 60 (28) |
| **Clinical presentation at enrolment** |  |  |  |  |
| Respiratory distress | 42 (19) | 57 (27) | 54 (25) | 45 (21) |
| Oxygenation | 7 (3.2) | 7 (3.3) | 9 (4.2) | 5 (2.4) |
| Impaired circulation | 7 (3.2) | 8 (3.8) | 6 (2.8) | 9 (4.3) |
| Impaired conscious level | 13 (6.0) | 26 (12) | 19 (8.8) | 20 (9.4) |
| Pulse > 180 per min | 35 (16) | 32 (15) | 33 (15) | 34 (16) |
| Haemoglobin < 7g/dl | 14 (6.5) | 22 (10) | 18 (8.3) | 18 (8.5) |
| Blood glucose < 3mmol/L | 16 (7.4) | 12 (5.6) | 8 (3.7) | 20 (9.4) |
| WBC count < 4 or > 17.5 x 10<sup>9</sup>/L | 65 (30) | 64 (30) | 68 (31) | 61 (29) |
| Temperature <36 or >38.5°C | 58 (27) | 63 (30) | 68 (31) | 53 (25) |
| Very low MUAC | 181 (84) | 176 (83) | 191 (88) | 166 (78) |
| Axillary temperature |  |  |  |  |
| <36°C | 18 (8.3) | 13 (6.1) | 20 (9.2) | 11 (5.2) |
| 36 to 37.5°C | 158 (73) | 150 (70) | 149 (69) | 159 (75) |
| >37.5°C | 40 (19) | 50 (24) | 48 (22) | 42 (20) |
| Respiratory rate |  |  |  |  |
| Bradypnoea | 20 (9.3) | 20 (9.4) | 23 (11) | 17 (8.0) |
| Normal | 102 (47) | 87 (41) | 94 (43) | 95 (45) |
| Tachypnoea | 94 (44) | 106 (50) | 100 (46) | 100 (47) |
| Capillary refill |  |  |  |  |
| <2s | 129 (60) | 124 (58) | 128 (59) | 125 (59) |
| 2 to 3s | 80 (37) | 81 (38) | 83 (38) | 78 (37) |
| >3s | 7 (3.2) | 8 (3.8) | 6 (2.8) | 9 (4.3) |
| Conscious level |  |  |  |  |
| Alert | 202 (94) | 187 (88) | 197 (91) | 192 (91) |
| Non-alert | 14 (6.5) | 26 (12) | 20 (9.2) | 20 (9.4) |
| Sunken eyes | 40 (19) | 49 (23) | 40 (18) | 49 (23) |
| **Syndrome/bedside investigations** |  |  |  |  |
| Gastroenteritis | 127 (59) | 119 (56) | 124 (57) | 122 (58) |
| Severe pneumonia | 37 (17) | 41 (19) | 41 (19) | 37 (17) |
| Sepsis diagnosis by clinician | 62 (29) | 64 (30) | 69 (32) | 57 (27) |
| Malaria RDT positive | 16 (7.4) | 23 (11) | 18 (8.3) | 21 (9.9) |
| HIV status |  |  |  |  |
| Negative | 168 (78) | 157 (74) | 164 (76) | 161 (76) |
| Infected | 18 (8.3) | 21 (9.9) | 17 (7.8) | 22 (10) |
| Unknown | 30 (14) | 35 (16) | 36 (17) | 29 (14) |
| Cerebral palsy, neurological problem or epilepsy | 26 (12) | 25 (12) | 32 (15) | 19 (9.0) |
| **Caregiver Characteristics** |  |  |  |  |
| Biological mother HIV status |  |  |  |  |
| Reactive | 10 (4.6) | 10 (4.7) | 11 (5.1) | 9 (4.3) |
| Known positive | 25 (12) | 23 (11) | 21 (9.7) | 27 (13) |
| Non-reactive | 140 (65) | 136 (64) | 143 (66) | 133 (63) |
| Unknown | 41 (19) | 44 (21) | 42 (19) | 43 (20) |

IQR; Interquartile Range, Small birth size; Low birth weight or born preterm, sd; standard deviation, MUAC; Mid-upper arm circumference in cm, WHZ; Weight-for-Height Z score, WAZ; Weight-for-age z score, HAZ; Height-for-age z score, WBC; While blood cells, normal respiratory rate; respiratory rate/min 30–60 for those aged <12months, 24–40 for those aged 12 to <36 months and 22–34 for those aged ≥36 months, bradypnoea; respiratory rate/min <30 for those aged <12months, <24 for those aged 12–<36 months and <22 for those aged ≥36 months, tachypnoea; respiratory rate/min >60 for those aged <12 months, >40 for those aged 12–<36 months and >34 for those aged ≥36 months.

### Follow up, mortality and retention

The children were in follow-up for 733.6 child-months during which 71/429 (16.6%) died; mortality rate of 96.8 (95% CI 76.7 122.1) deaths per 1000 child-months. The median (IQR) time to death was 8 (3 19) days. Overall, 340 of surviving children completed study follow-up and 18 (4.2%) had incomplete follow up. Incomplete follow up comprised 14 voluntary withdrawals, three lost-to-follow-up and one protocol withdrawal because of meeting an exclusion criterion (**Fig 1**). Participant flow chart and outcome data are also presented stratified by the four groups in **S3 Table and S1 Fig**.

**Figure 1.**
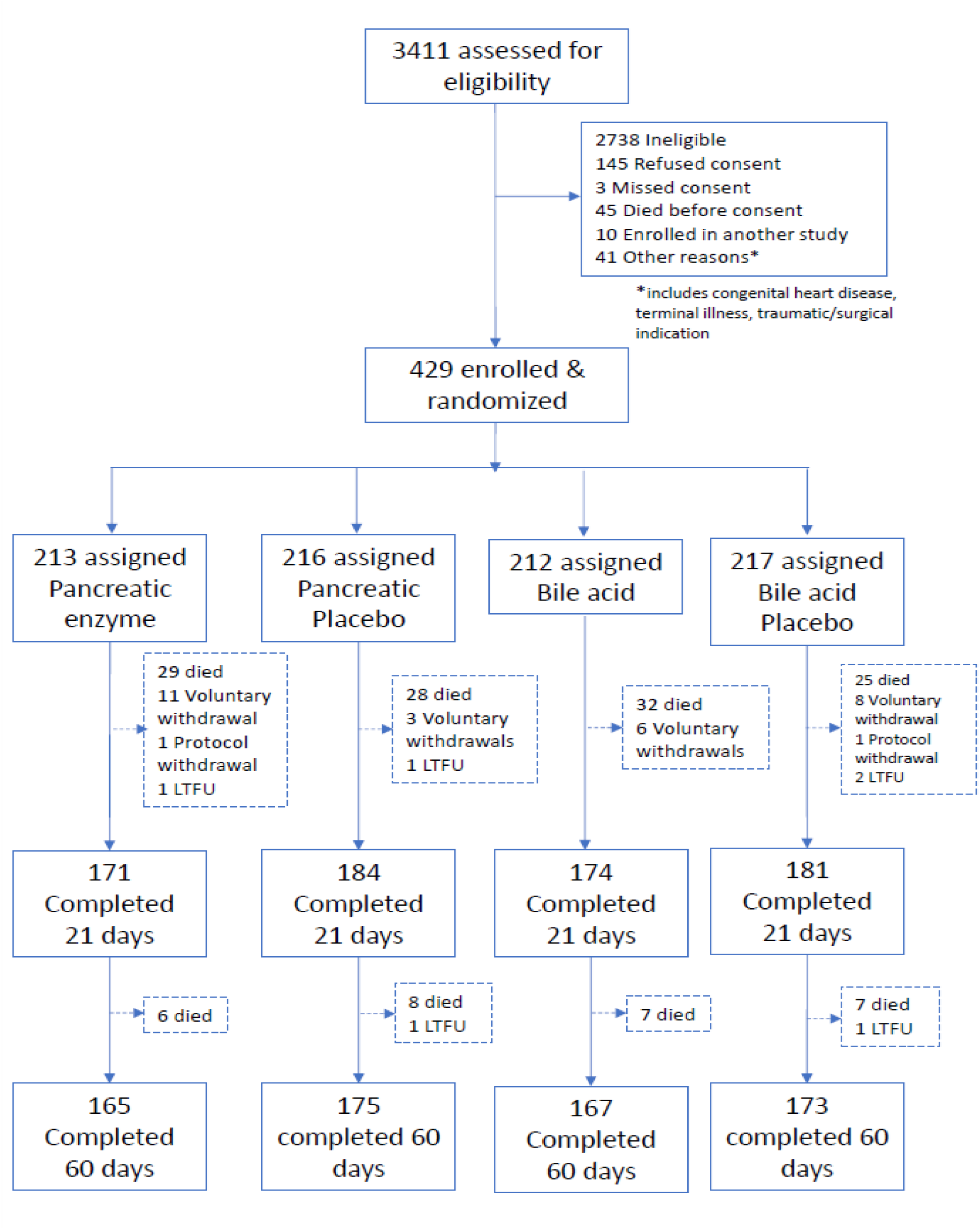
Trial participant flow chart.

### Treatment fidelity

Overall, 355/429 (83%) children completed 21 days receiving the interventions: 171/213 (80%) and 184/216 (85%) of children assigned PE and PE-placebo, respectively, and 174/212 (82%) and 181/217 (83%) assigned to BA and BA-placebo respectively (**Fig1**).

### Interim analyses

Both PE and BA interventions were retained after the first interim analysis, as their posterior probability of non-inferiority exceeded the pre-defined 80% threshold. However, both interventions were discontinued after the second interim analyses because their posterior probability of superiority fell below the pre-defined threshold of 0.8 (80%). Based on these results, the DMSB recommended stopping the trial, a decision that was adopted by the Trial Steering Committee.

### Primary outcome

A total of 71 deaths (16.6%) occurred within 60 days, with 57 (80%) of these occurring within 21 days, during the period of trial product use. Among the 213 children allocated to PE, 35 (16%) died versus 36/216 (17%) in those given PE-placebo, irrespective of receiving BA or BA-placebo (HR 1.02 (95% CI 0.64 1.62), log-rank p-value=0.94; **Table 2**, **Fig 1** and **Fig 2a**). Adjusting for recruiting site did not alter this finding (p-value=0.92). There was no evidence of interaction with the BA allocation (p-value=0.58). Among children allocated to BA, 39 (18%) died compared to 32 (15%) children receiving BA-placebo, irrespective of PE or PE-placebo allocation (HR 1.27; 95% CI: 0.79 2.02, log-rank p-value=0.32) (**Table 2**, **Fig 1** and **Fig 2b)**.

**Table 2.** Trial outcomes.

|  | Pancreatic enzymes randomization |  | Bile acid randomization |  |
| --- | --- | --- | --- | --- |
|  | Placebo (n=216) | Pancreatic enzymes (n=213) | Placebo (n=217) | Bile acid (n=212) |
| **Primary Outcome** |  |  |  |  |
| All-cause deaths |  |  |  |  |
| N (%) | 36 (17) | 35 (16) | 32 (15) | 39 (18) |
| Mortality rate (95% CI)/1000 CM | 95.1 (68.6-132) | 98.6 (70.8-137) | 85.0 (60.1-120) | 109.2 (79.8-150) |
| Mortality rate difference (95% CI) | 3.47 (-41.6 to 48.5) |  | 24.2 (-20.9 to 69.4) |  |
| Hazard ratio (95% CI) | Reference | 1.02 (0.64-1.62) | Reference | 1.27 (0.79-2.02) |
| **Secondary Outcomes** |  |  |  |  |
| Hospital readmissions after index discharge |  |  |  |  |
| Episodes | 23 | 21 | 18 | 26 |
| Incidence rate (95% CI)/1000 CM | 78.7 (52.3-118) | 75.9 (49.5-117) | 62.0 (39.1-98.4) | 93.4 (63.6-137) |
| Incidence rate difference (95%CI) | -2.7 (-48.2 to 43.0) |  | 31.4 (-14.5 to 77.3) |  |
| Hazard ratio (95% CI) | Reference | 0.97 (0.54-1.75) | Reference | 1.51 (0.83-2.75) |
| All SAEs |  |  |  |  |
| Episodes | 76 | 77 | 74 | 79 |
| Incidence rate (95% CI)/1000 CM | 230 (184-288) | 247 (197-308) | 225 (179-282) | 252 (202-314) |
| Incidence rate difference (95%CI) | 16.6 (-58.9 to 92.2) |  | 27.2 (-48.4 to 102.8) |  |
| Hazard ratio (95% CI) | Reference | 1.08 (0.79-1.49) | Reference | 1.09 (0.79-1.50) |
| Grade 3 & 4 toxicity* |  |  |  |  |
| Episodes | 2 | 2 | 2 | 2 |
| Incidence rate (95% CI) /1000 CM | 14.9 (3.74-59.7) | 15.6 (3.91-62.5) | 15.0 (3.74-59.8) | 15.6 (3.91-62.5) |
| Incidence rate difference (95%CI) | 0.7 (-29.3 to 30.7) |  | 0.7 (-29.3 to 30.6) |  |
| Hazard ratio (95% CI) | Reference | 1.04 (0.15-7.38) | Reference | 1.04 (0.15-7.36) |
| Days spent in hospital, median (IQR) | 8 (5-13) | 7 (5-13) | 8 (5-14) | 7 (5-12) |
| Regression coefficient (95% CI) | Reference | -0.03 (-0.14 to 0.09) | Reference | -0.07 (-0.18-0.05) |
| Days with sepsis | 255 | 308 | 312 | 251 |
| Regression coefficient (95% CI)# | Reference | 0.16 (-0.12 to 0.43) | Reference | -0.15 (-0.42 to 0.13) |
| Days with diarrhea | 567 | 556 | 532 | 591 |
| Regression coefficient (95% CI)# | Reference | 0.07 (-0.10 to 0.23) | Reference | 0.07 (-0.09 to 0.24) |
| Days on second line antibiotics | 794 | 715 | 832 | 677 |
| Regression coefficient (95% CI)# | Reference | -0.03 (-0.20 to 0.14) | Reference | -0.23 (-0.40 to -0.06) |
| Days on third line antibiotics | 126 | 72 | 99 | 99 |
| Regression coefficient (95% CI)# | Reference | -0.23 (-0.60 to 0.13) | Reference | -0.09 (-0.46 to 0.27) |

CM; Child-months, SAEs; Severe Adverse Events, IQR; Interquartile Range, \*toxicity events during 21 days from enrolment when receiving study intervention, # Regression coefficient from zero inflated negative binomial regression models.

**Figure 2:**
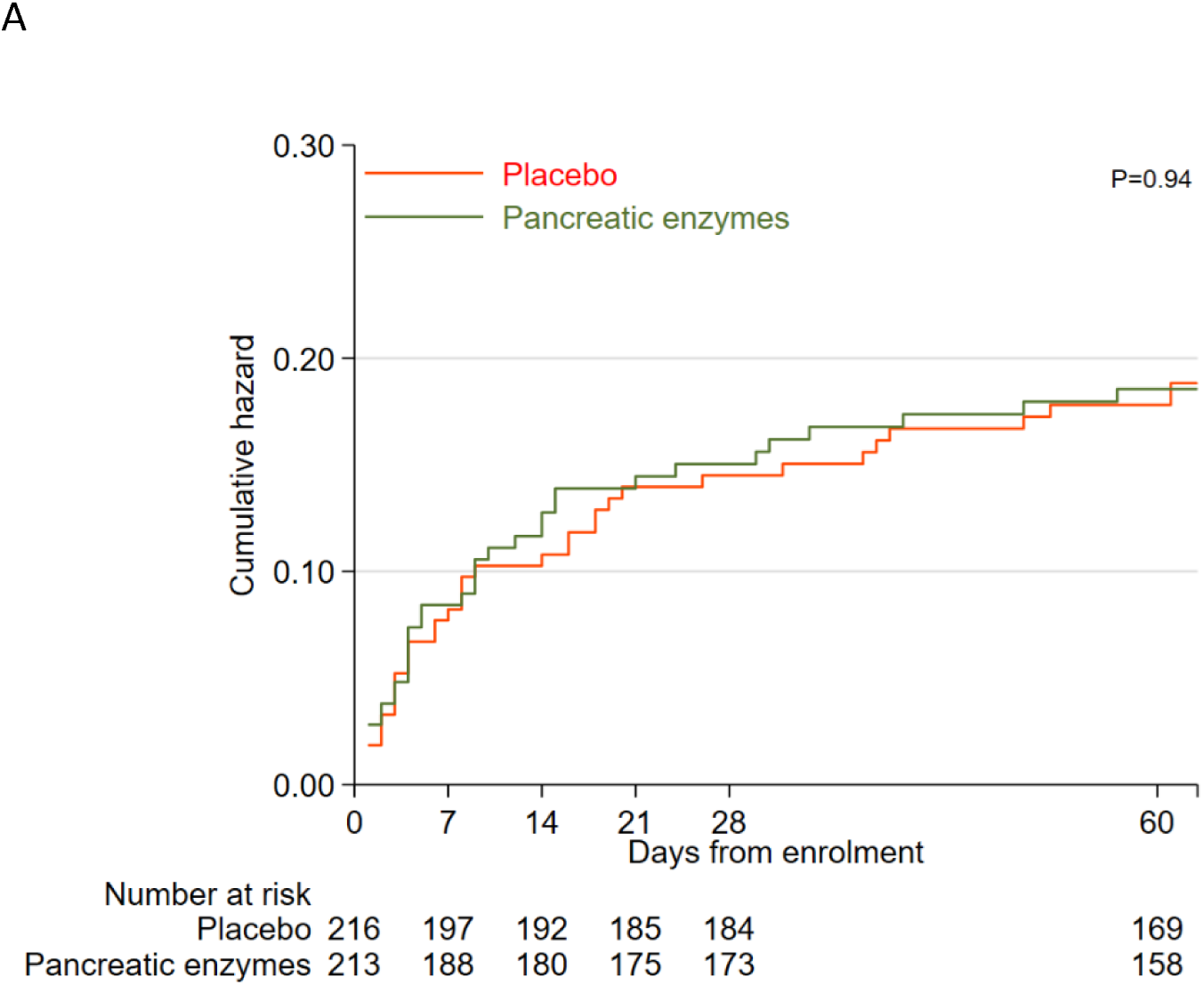
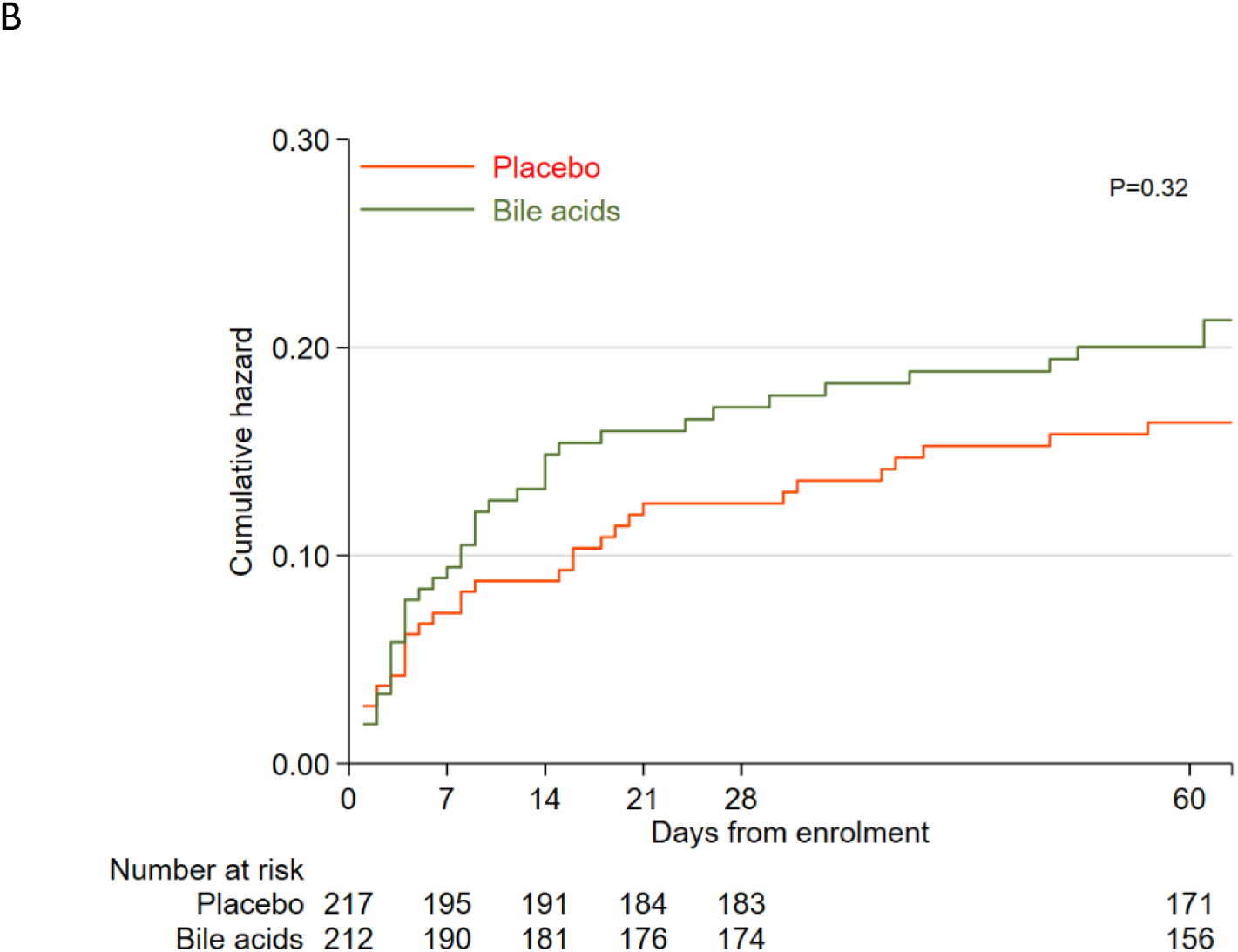
Mortality from enrolment to 60 days: A) among children randomized to pancreatic enzymes versus placebo and B) among children randomized to bile-acid versus placebo.

Again, there was no evidence of interaction with PE assignment (p-value=0.95) and adjustment for site did not impact results (p-value= 0.33). Deaths stratified by the four treatment allocations are shown in **S3 Table.** Neither PE nor BA significantly altered mortality within the first 21 days (**S4 Table**). Assigned causes of deaths are shown in **S5 Table.**

### SAEs

A total of 153 SAEs occurred among 138/429 (32%) children: these included 77 events in children receiving PE versus 76 in the PE-placebo group (HR 1.08; 95% CI: 0.79 1.49); while 79 events occurred in children receiving BA versus 74 in the BA-placebo group (HR 1.09; 95% CI: 0.79 1.50) (**Table 2**). The most common SAEs were due to prolonged or re-hospitalization with gastroenteritis (n=62/153, 41%), severe pneumonia (n=60/153, 39%) and sepsis (n=56/153, 37%) **S6 Table**. There were 44 hospital readmissions among 37 children of the 372 children discharged alive and followed up (seven children were readmitted twice): This included 21 readmissions in the PE group versus 23 in the PE-placebo group (HR 0.97; 95% CI: 0.54 1.75); and 26 in the BA group versus 18 in the BA-placebo group (HR 1.51; 95% CI: 0.83 2.75) (**Table 2**).

### Prespecified toxicities

During the 21 days of intervention, grade 3 or 4 toxicity was reported among four children. One developed hepatic encephalopathy with unknown diagnosis primary liver disease and discontinued the trial interventions after 16 days (32 doses) of ‘PE-placebo and BA’. Two children experienced diarrhea with hypotensive shock (one receiving PE and BA-placebo and the other PE and BA). The fourth child was receiving PE and BA-placebo and experienced a marked increase in stool frequency (≥7 stools in 24 hours) (**Table 2** & **S7 Table**). None of the toxicities were determined to be attributable to study interventions.

### Clinical secondary outcomes

Diarrhea and use of second-line antibiotics were common during the index admission. Of all the clinical secondary outcomes, one reached statistical significance: children allocated to BA spent fewer days on second-line antibiotics than children receiving BA-placebo (677 vs. 832 days, incidence rate ratio 0.78 (95% CI: 0.65 0.95), p-value=0.01) (**Table 2**). Other secondary outcomes including days spent in hospital and days with sepsis, diarrhea and use of third-line antibiotics did not significantly differ by randomized allocation.

### Growth

Overall, MUAC, WHZ and WAZ increased between enrollment and day 60, but HAZ declined (**S2 Fig**). There was no evidence that changes in MUAC, WHZ and WAZ from enrollment to day 21 or day 60 differed between allocation groups (all p-values ≥0.05, **Table 3** and **S2 Fig**).

**Table 3.**
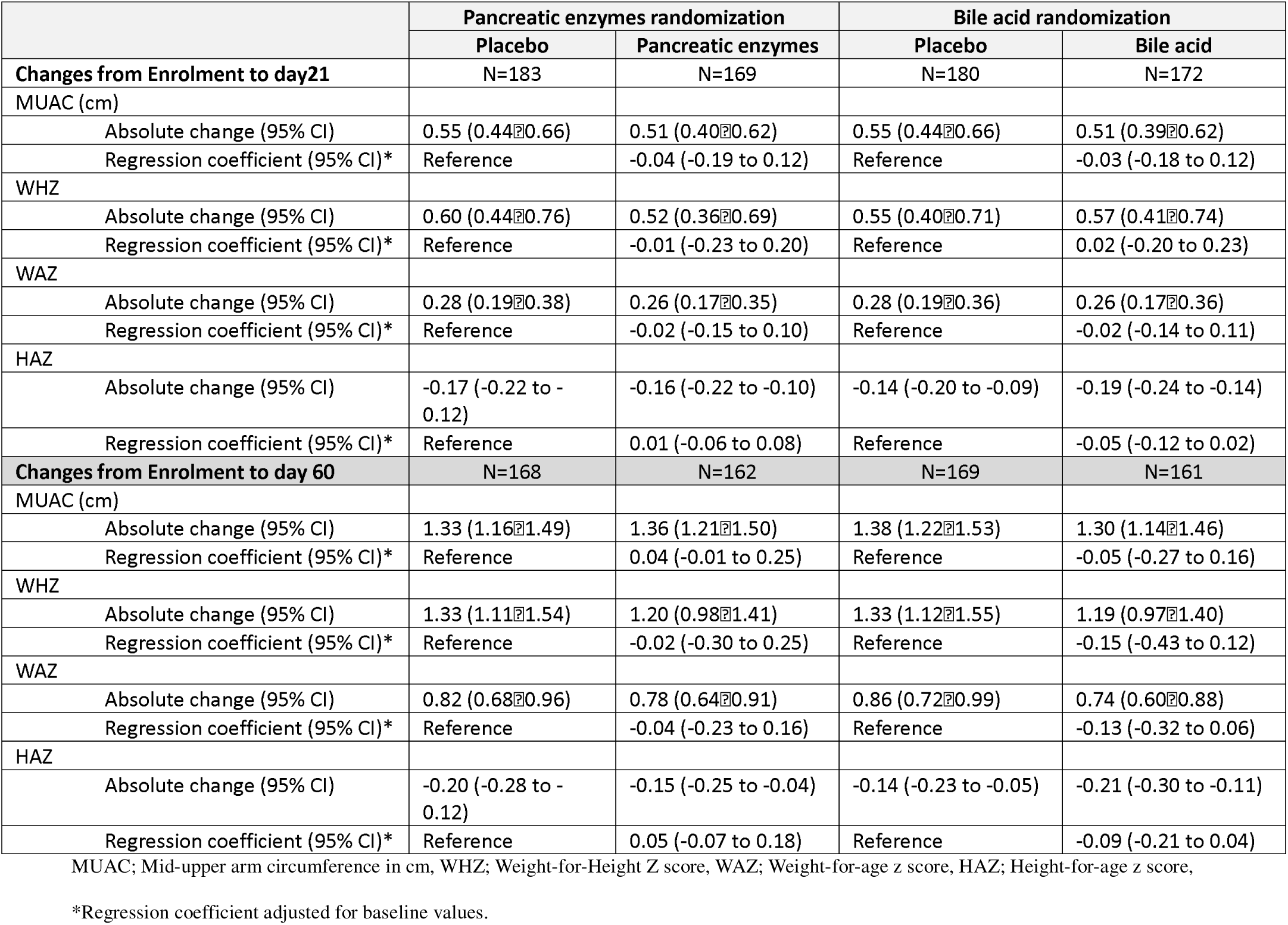
Growth from enrolment to day 21 and day 60.

### Adherence

Adherence to both study formulations was moderate to high across treatment groups, with estimated mean adherence ranging from approximately 60% to 84% based on weight differences of returned bottles (**S8 Table**). Using beta regression, adherence index was lower by 9% in the pancreatic-enzyme group compared to placebo (p = 0.017), while no difference was observed between bile-acid and placebo formulations (p = 0.76). Adherence data were incomplete for a subset of participants due to lost-to-follow up after discharge (n=17, 5.1%), missing bottle weight at discharge (PE, n=30 (9.0%); Urso, n=13 (3.9%)) or unreturned bottles discharge (PE, n=11 (6.5%); Urso, n=8 (4.9%)).

## Discussion

Mortality during hospitalization in acutely ill children with severe malnutrition is more than five times higher than those without severe malnutrition ^2^. We tested the hypothesis that treating acutely ill severely malnourished children with pancreatic enzymes and/or bile acids would reduce mortality. The trial was stopped for futility after an interim analysis showed that neither intervention or their combination was likely to demonstrate superiority over placebo. We also observed no meaningful differences in SAE, clinical secondary outcomes or growth, with the exception of children who received treatment with BA spent fewer days on second-line antibiotics compared to those receiving BA-placebo.

Sepsis, pneumonia, gastroenteritis, and dehydration are commonly associated with mortality in children hospitalized with acute illness and severe malnutrition. Severely ill malnourished children exhibit profound disruptions in energy metabolism that affects cellular processes and likely impairs their capacity to mount an effective immune response to infections ^26^. Translocation of bacterial components and viable bacteria from the gastrointestinal tract to systemic circulation is described in both critical illness and severe malnutrition ^8,27,28^. Recent data suggest bacterial overgrowth in the small intestine is common in malnourished children ^29^.

Both pancreatic enzymes and bile acids have direct bacteriocidic properties ^9,10,30,31^. Bile acids can disrupt bacterial membranes, induce oxidative stress and cause DNA damage, thereby reducing bacterial viability ^12,31,32^. In addition, pancreatic enzymes and bile acids have been demonstrated to modulate the gut microbiome ^32^. In murine models, pancreatic enzyme treatment increased the abundance of ‘beneficial’ intestinal bacteria, such as *Akkermansia muciniphila* ^33^. Ursodeoxycholic acid, commonly used to treat patients with cholestatic diseases, reduced intestinal dysbiosis and intestinal inflammation in a murine model of inflammatory bowel disease ^34^. Bile acids have been shown to improve intestinal barrier function in disease models ^35^. Although a pilot trial conducted in 2014 suggested a possible benefit on survival from pancreatic enzyme supplementation, its findings were limited since the study was unblinded and not powered for mortality ^20^. In contrast, our current randomized, placebo-controlled trial was specifically powered to evaluate mortality as a primary outcome in a severely ill population. The mortality point estimate for pancreatic enzyme treatment does not suggest inadequate power, but rather an absence of therapeutic effect. While bile acids reduced the number of days on second-line antibiotics, this intervention had a point estimate for mortality in favor of placebo, raising potential safety concerns when administered during the critical phase of illness in this population.

The lack of mortality benefit from pancreatic enzyme or bile acids should be considered within the physiological context of critical illness and severe malnutrition. During early hospitalization, malnourished children often experience sepsis, hypovolemia, gastrointestinal and metabolic dysfunction. In our trial, over 50% of deaths occurred within 8 days, at time when the physiological response to interventions is likely blunted due to significant multi-organ dysfunctions. The proposed mechanism of action for pancreatic enzymes and bile acids, such as enhancing nutrient absorption, supressing bacterial overgrowth, and improving gut barrier integrity may not yield immediate survival benefits. Such effects likely require both a longer treatment duration and restoration of metabolic and energy homeostasis. It remains plausible that these interventions could support growth or recovery in less acutely ill children with severe malnutrition, and further investigation in that context may be warranted.

A key strength of our study is its multi-national design, conducted across diverse geographies where malnutrition is prevalent, thereby increasing the generalizability of the trial findings. Being under the umbrella of the CHAIN Network ensured a harmonized, high-quality trial platform, leveraging over seven years of collaborative experience across sites. Doses used for the pancreatic enzymes and ursodeoxycholic acid were similar to that prescribed to children with exocrine pancreas insufficiency or cholestatic disease. However, we were unable to directly observe adherence to these treatments at home. The estimates of adherence should be interpreted cautiously. These data were incomplete for a subset of participants which may introduce bias. Moreover, estimations relied on change in bottle weight which assumes that doses were dispensed and consumed as intended. In practice, spillage, sharing, or loss of liquid during administration likely occurred, leading to systematic overestimation of adherence. Also, no biological or pharmacokinetic marker was available to validate drug ingestion, so the adherence estimates reflect reported dosing behavior rather than confirmed exposure. In addition, we were unable to assess whether the interventions reduced small intestinal bacterial overgrowth, altered microbiome composition or modified the bile acid pool. Having more proximal mechanistic outcomes may have identified more subtle biological effects that were not reflected in outcomes of mortality, SAEs, or growth over 60 days.

## Conclusions

These data indicate that empiric treatment of acutely ill severely malnourished children with pancreatic enzymes and/or ursodeoxycholic acid does not reduce mortality at 60 days. The use of an adaptive trial design provide an efficient and cost-effective approach to reach this endpoint. Our work contributes to a growing body of intervention studies performed over the last decade that have trialed various antibiotics or nutritional modifications, and were unable to demonstrate a survival benefit in ill malnourished children. This illustrates the complex and multifactorial pathophysiology of severe malnutrition in the context of acute illness. Improving outcomes will require the targeting of several pathways, potentially in a sequential manner that aligns with key phases of recovery. This study did not reveal supporting evidence that BA, PE or their combination substantially alter the clinical course of malnourished children in hospital or after discharge. Future progress will likely depend on novel interventions arising from a more holistic approach, leading to a package of interventions that may include medical, social and financial domains.

## Data Availability

All data produced in the present study are available upon reasonable request to the authors

## Acknowledgements

We are deeply grateful to the patients and their caregivers for participating in the study. We also thank investigators and support staff at all participating centers for their dedication. We acknowledge the valuable oversight provided by the DSMB members Dr. Rajiv Bahl, Dr. Ann Strode, Dr. Charles Opondo and Dr. Naor Bar Seev. We acknowledge also the support of Trial Steering Committee members Dr. Indi Trehan, Dr. Marko Kerac and Dr. Beatrice Amadi. This study was supported by the Bill and Melinda Gates Foundation (INV-000791). For the purpose of open access, the authors have applied a CC BY public copyright license to any author-accepted manuscript version arising from this submission.

## Author Contributions

**Conceptualization:** Robert H.J. Bandsma, Wieger P. Voskuijl, Mohammod Jobayer Chisti, Tahmeed Ahmed, Judd L. Walson, James A. Berkley

**Data curation:** Moses Ngari, Narshion Ngao, James A Berkley

**Formal analysis:** Moses Ngari, Alex Dmitrienko, Robert Bandsma, Judd Walson, James A Berkley

**Funding acquisition**: Robert H.J. Bandsma, Judd L. Walson, James A. Berkley

**Investigation:** Mohammod Jobayer Chisti, Emmie Mbale, Abu Sadat Mohammad Sayeem Bin Shahid, Ezekiel Mupere, Christina L Lancioni, Christopher Marong, Christopher Lwanga, Michael Atuhairwe, Chisomo Eneya, Kirkby D Tickell, Caroline Tigoi, Moses Mburu

**Methodology:** Robert H.J. Bandsma, Wieger P. Voskuijl, Mohammod Jobayer Chisti, Moses Ngari, Narshion Ngao, Chikondi Makwinja, Abu Sadat Mohammad Sayeem Bin Shahid, Johnstone Thitiri, Judd L. Walson, James A. Berkley

**Project administration:** Robert H.J. Bandsma, Dennis Chasweka, Johnstone Thitiri, Isaiah Njagi, Judd L. Walson, James A. Berkley

**Supervision:** Mohammod Jobayer Chisti, Emmie Mbale, Ezekiel Mupere, Christina L Lancioni, Christopher Lwanga, Kirkby D Tickell, Isaiah Njagi, Johnstone Thitiri, Caroline Tigoi, Robert H.J. Bandsma, Wieger P. Voskuijl, Tahmeed Ahmed, Judd L. Walson, James A. Berkley

**Writing - original draft:** Robert H.J. Bandsma, Wieger P. Voskuijl, Mohammod Jobayer Chisti, Moses Ngari, Tahmeed Ahmed, Judd L. Walson, James A. Berkley

**Writing – review and editing:** Robert H. J. Bandsma, Mohammod Jobayer Chisti, Wieger P. Voskuijl, Moses M. Ngari, Ezekiel Mupere, Isaiah Njagi, Johnstone Thitiri, Emmie Mbale, Isabel Potani, Dennis Chasweka, Chikondi Makwinja, Chisomo Eneya, Celine Bourdon, Abu Sadat Mohammad Sayeem Bin Shahid, Gazi Md. Salahuddin Mamun, Christina L. Lancioni, Christopher Marong, Christopher Lwanga, Michael Atuhairwe, Kirkby D. Tickell, Caroline Tigoi, Moses Mburu, Narshion Ngao, Alex Dmitrienko, Tahmeed Ahmed, Judd L. Walson, James A. Berkley

## Supplementary Materials

### Supplementary Results

**S1Table.**
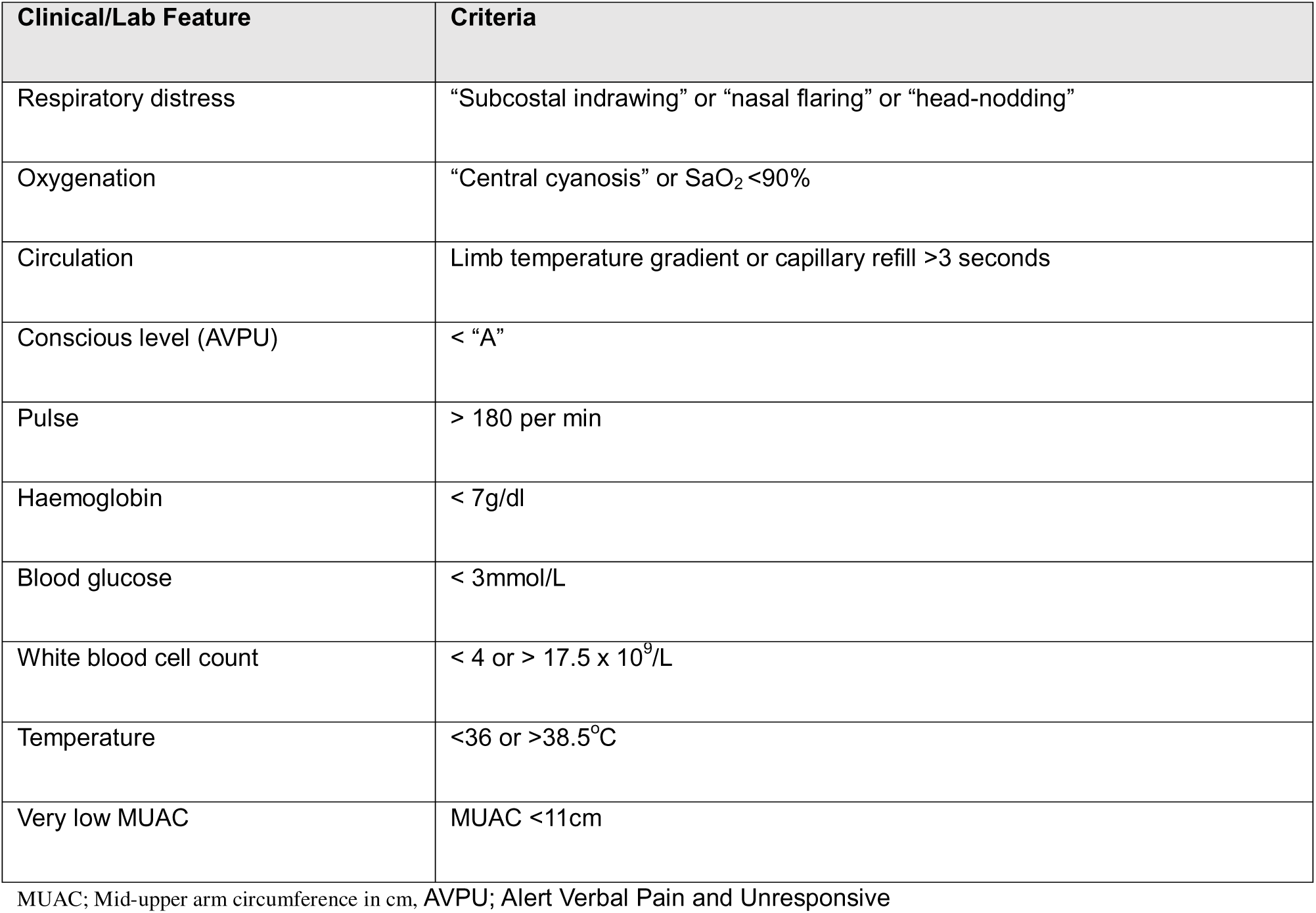
Severity features, used as inclusion criteria with two or more required for enrolment.

Supplementary Results S1Table. Severity features, used as inclusion criteria with two or more required for enrolment.
| Clinical/Lab Feature | Criteria |
| --- | --- |
| Respiratory distress | “Subcostal indrawing” or “nasal flaring” or “head-nodding” |
| Oxygenation | “Central cyanosis” or $\text{SaO}_2 < 90\%$ |
| Circulation | Limb temperature gradient or capillary refill $> 3$ seconds |
| Conscious level (AVPU) | $< \text{“A”}$ |
| Pulse | $> 180$ per min |
| Haemoglobin | $< 7\text{g/dl}$ |
| Blood glucose | $< 3\text{mmol/L}$ |
| White blood cell count | $< 4$ or $> 17.5 \times 10^9/\text{L}$ |
| Temperature | $< 36$ or $> 38.5^\circ\text{C}$ |
| Very low MUAC | MUAC $< 11\text{cm}$ |

MUAC; Mid-upper arm circumference in cm, AVPU; Alert Verbal Pain and Unresponsive

**S2 Table.** Participant baseline characteristics by the four treatment allocations.

| Characteristics | Randomization arm |  |  |  |
| --- | --- | --- | --- | --- |
|  | Pancreatic enzymes only (n=108) | Bile acid only (n=107) | Pancreatic enzymes + Bile acid (n=105) | Double placebo (n=109) |
| Age in months; median (IQR) | 9.8 (4.5–18.2) | 10.6 (5.8–16.9) | 10.0 (4.6–21.0) | 10.4 (4.8–16.2) |
| Sex |  |  |  |  |
| Female | 55 (51) | 44 (41) | 49 (47) | 47 (43) |
| Small birth size | 30 (28) | 31 (29) | 36 (34) | 33 (30) |
| Previous hospital admission | 38 (35) | 44 (41) | 38 (36) | 34 (31) |
| Reported antibiotics 7 days prior to admission | 52 (48) | 47 (44) | 37 (35) | 42 (39) |
| Currently breastfeeding | 55 (51) | 40 (37) | 37 (35) | 49 (45) |
| Attending outpatient nutrition program prior to admission |  |  |  |  |
| No | 94 (87) | 90 (84) | 90 (86) | 96 (88) |
| Supplementary/Therapeutic | 14 (13) | 17 (16) | 15 (14) | 13 (12) |
| Site |  |  |  |  |
| Blantyre, Malawi | 31 (29) | 30 (28) | 29 (28) | 29 (27) |
| Dhaka, Bangladesh | 30 (28) | 29 (27) | 28 (27) | 30 (28) |
| Kampala, Uganda | 20 (19) | 20 (19) | 21 (20) | 23 (21) |
| Kilifi, Kenya | 23 (21) | 22 (21) | 25 (24) | 24 (22) |
| Nairobi, Kenya | 4 (3.7) | 6 (5.6) | 2 (1.9) | 3 (2.8) |
| Anthropometry: mean ±sd | | | | |
| MUAC | 10.3 ±1.15 | 10.4 ±1.38 | 10.3 ±1.32 | 10.42±0.84 |
| WHZ | -3.29 ±1.28 | -3.57 ±1.24 | -3.49 ±1.54 | -3.42 ±1.46 |
| WAZ | -4.55 ±0.92 | -4.70 ±1.11 | -4.71 ±1.23 | -4.44 ±1.18 |
| HAZ | -3.65 ±1.48 | -3.76 ±1.79 | -3.59 ±1.82 | -3.42 ±1.88 |
| Oedema | 20 (19) | 31 (29) | 29 (28) | 23 (21) |
| **Clinical presentation at enrolment** |  |  |  |  |
| Respiratory distress | 30 (28) | 18 (17) | 27 (26) | 24 (22) |
| Oxygenation | 3 (2.8) | 2 (1.9) | 3 (2.9) | 6 (5.5) |
| Impaired circulation | 3 (2.8) | 4 (3.7) | 5 (4.8) | 3 (2.8) |
| Impaired conscious level | 8 (7.4) | 5 (4.7) | 16 (15) | 10 (9.2) |
| Pulse > 180 per min | 19 (18) | 21 (20) | 13 (12) | 14 (13) |
| Haemoglobin < 7g/dl | 13 (12) | 9 (8.4) | 9 (8.6) | 5 (4.6) |
| Blood glucose < 3mmol/L | 5 (4.6) | 13 (12) | 7 (6.7) | 3 (2.8) |
| WBC count < 4 or > 17.5 x 10<sup>9</sup>/L | 35 (32) | 32 (30) | 29 (28) | 33 (30) |
| Temperature <36 or >38.5°C | 30 (28) | 26 (24) | 33 (31) | 32 (29) |
| Very low MUAC | 91 (85) | 81 (76) | 85 (81) | 100 (93) |
| Axillary temperature |  |  |  |  |
| <36°C | 8 (7.4) | 6 (5.6) | 5 (4.8) | 12 (11) |
| 36 to 37.5°C | 75 (69) | 84 (79) | 75 (71) | 74 (68) |
| >37.5°C | 25 (23) | 17 (16) | 25 (24) | 23 (21) |
| Respiratory rate |  |  |  |  |
| Bradypnoea | 11 (10) | 8 (7.5) | 9 (8.6) | 12 (11) |
| Normal | 47 (44) | 55 (51) | 40 (38) | 47 (43) |
| Tachypnoea | 50 (46) | 44 (41) | 56 (53) | 50 (46) |
| Capillary refill |  |  |  |  |
| <2s | 69 (64) | 70 (65) | 55 (52) | 59 (54) |
| 2 to 3s | 36 (33) | 33 (31) | 45 (43) | 47 (43) |
| >3s | 3 (2.8) | 4 (3.7) | 5 (4.8) | 3 (2.8) |
| Conscious level |  |  |  |  |
| Alert | 99 (92) | 104 (97) | 88 (84) | 98 (90) |
| Non-alert | 9 (8.3) | 3 (2.8) | 17 (16) | 11 (10) |
| Sunken eyes | 17 (16) | 17 (16) | 32 (30) | 23 (21) |
| **Syndrome/bedside investigations** |  |  |  |  |
| Gastroenteritis | 60 (56) | 63 (59) | 59 (56) | 64 (59) |
| Severe pneumonia | 21 (19) | 17 (16) | 20 (19) | 20 (18) |
| Sepsis diagnosis by clinician | 33 (31) | 26 (24) | 31 (30) | 36 (33) |
| Malaria RDT positive | 10 (9.3) | 8 (7.5) | 13 (12) | 8 (7.3) |
| HIV status |  |  |  |  |
| Negative | 81 (75) | 85 (79) | 76 (72) | 83 (76) |
| Infected | 8 (7.4) | 9 (8.4) | 13 (12) | 9 (8.3) |
| Unknown | 19 (18) | 13 (12) | 16 (15) | 17 (16) |
| Cerebral palsy, neurological problem or epilepsy | 16 (15) | 10 (9.4) | 9 (8.6) | 16 (15) |
| **Caregiver Characteristics** |  |  |  |  |
| Biological mother HIV status |  |  |  |  |
| Reactive | 5 (4.6) | 4 (3.7) | 5 (4.8) | 6 (5.5) |
| Known positive | 9 (8.3) | 13 (12) | 14 (13) | 12 (11) |
| Non-reactive | 73 (68) | 70 (65) | 63 (60) | 70 (64) |
| Unknown | 21 (19) | 20 (19) | 23 (22) | 21 (19) |

**S3 Table.**
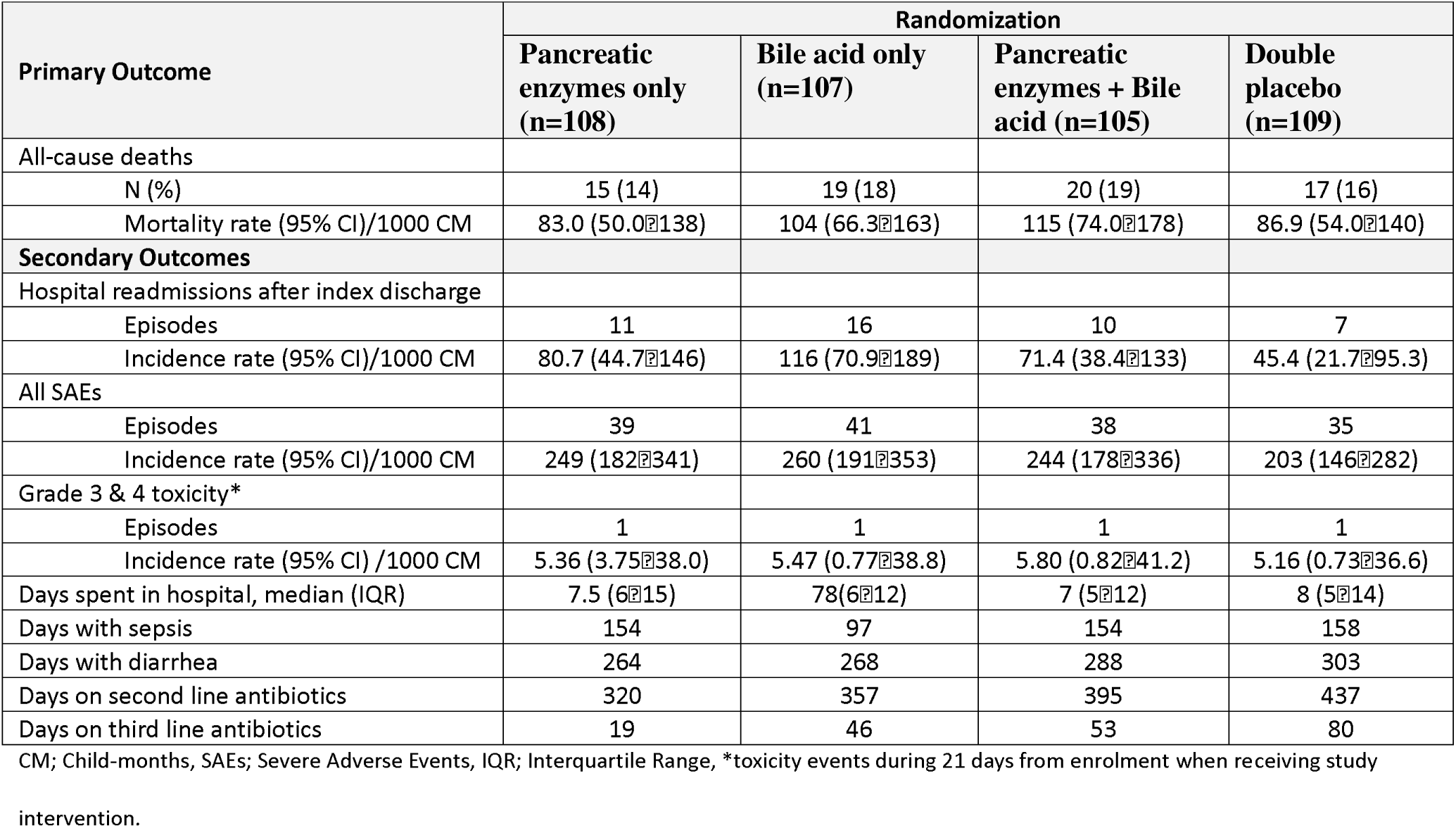
Trial outcomes stratified by the four treatment allocations.

**S4 Table.** Trial outcomes to 21 days.

|  | Pancreatic enzymes randomization |  | Bile acid randomization |  |
| --- | --- | --- | --- | --- |
|  | Placebo (n=216) | Pancreatic enzymes (n=213) | Placebo (n=217) | Bile acid (n=212) |
| All-cause deaths |  |  |  |  |
| Absolute numbers | 29 (13) | 28 (13) | 25 (12) | 32 (15) |
| Hazard ratio (95% CI) | Reference | 1.04 (0.62-1.76) | Reference | 1.28 (0.75-2.17) |
| **Secondary Outcomes** |  |  |  |  |
| All SAEs |  |  |  |  |
| Episodes | 51 | 53 | 50 | 54 |
| Hazard ratio (95% CI) | Reference | 1.09 (0.74-1.60) | Reference | 1.10 (0.75-1.61) |
| Grade 3 & 4 toxicity |  |  |  |  |
| Episodes | 2 | 2 | 2 | 2 |
| Hazard ratio (95% CI) | Reference | 1.04 (0.15-7.38) | Reference | 1.04 (0.15-7.36) |
| Days spent in hospital, median (IQR) | 8 (5-13) | 7 (5-13) | 8 (5-14) | 7 (5-12) |
| Regression coefficient (95% CI) | Reference | -0.03 (-0.14 to 0.09) | Reference | -0.07 (-0.18 to 0.05) |
| Days with sepsis | 233 | 308 | 290 | 251 |
| Regression coefficient (95% CI)<sup>#</sup> | Reference | 0.24 (-0.02 to 0.50) | Reference | -0.09 (-0.35 to 0.17) |
| Days with diarrhea | 560 | 555 | 526 | 589 |
| Regression coefficient (95% CI)<sup>#</sup> | Reference | 0.07 (-0.08 to 0.24) | Reference | 0.08 (-0.07 to 0.24) |
| Days on second line antibiotics | 776 | 711 | 814 | 673 |
| Regression coefficient (95% CI)<sup>#</sup> | Reference | -0.01 (-0.18 to 0.16) | Reference | -0.21 (-0.37 to -0.05) |
| Days on third line antibiotics | 126 | 72 | 99 | 99 |
| Regression coefficient (95% CI)<sup>#</sup> | Reference | -0.23 (-0.60 to 0.13) | Reference | -0.09 (-0.46 to 0.27) |

SAEs; Severe Adverse Events, IQR; Interquartile Range, <sup>#</sup>regression coefficient from zero inflated negative binomial regression models.

**S5 Table.** Assigned causes of deaths according to intervention groups.

| Cause of death* | Pancreatic enzymes randomization |  | Bile acid randomization |  |
| --- | --- | --- | --- | --- |
|  | Placebo (n=36) | Pancreatic enzymes (n=35) | Placebo (n=32) | Bile acid (n=39) |
| Severe pneumonia | 22 | 12 | 17 | 17 |
| Sepsis | 15 | 16 | 16 | 15 |
| Severe acute malnutrition | 11 | 10 | 11 | 10 |
| Gastroenteritis | 8 | 6 | 6 | 8 |
| Shock | 5 | 5 | 3 | 7 |
| HIV related | 3 | 4 | 2 | 5 |
| Meningitis | 3 | 3 | 2 | 4 |
| Severe anaemia | 4 | 0 | 2 | 2 |
| Malaria | 0 | 3 | 0 | 3 |
| TB | 2 | 0 | 0 | 2 |

\*Some children were assigned more than one causes of death.

**S6 Table.** List of Serious Adverse Event.

|  | Site | Pancreatic enzymes | Bile acids | Sex | Age in months | HIV test | Attribution | Outcome | Anaemia | Pneumonia | Pulmonary TB | Gastroenteritis | Sepsis | Malaria | Cerebral Palsy | Other Diagnosis |
| --- | --- | --- | --- | --- | --- | --- | --- | --- | --- | --- | --- | --- | --- | --- | --- | --- |
| 1 | Kilifi | No | No | Female | 6-10 | Negative | No | Resolved | No | Yes | No | No | No | No | No | Exacerbation of congenital cardiac disease |
| 2 | Kilifi | No | No | Male | 6-10 | Unknown | No | Resolved | No | No | No | Yes | No | No | No |  |
| 3 | Kilifi | Yes | Yes | Male | 0-5 | Infected | No | Resolved | No | No | No | Yes | No | No | No | Otitis media |
| 4 | Kilifi | Yes | Yes | Male | 0-5 | Infected | No | Resolved | No | No | No | Yes | Yes | No | No |  |
| 5 | Kilifi | Yes | Yes | Male | 6-10 | Unknown | Unlikely | Resolved | No | No | No | Yes | No | No | No |  |
| 6 | Kilifi | No | Yes | Male | 6-10 | Unknown | Unlikely | Fatal | No | Yes | No | No | No | No | No |  |
| 7 | Kilifi | No | Yes | Female | 11-15 | Unknown | Unlikely | Fatal | No | No | No | No | Yes | No | No | Osteomyelitis, other encephalopathy |
| 8 | Kilifi | Yes | Yes | Male | 16-20 | Unknown | No | Fatal | No | No | No | No | Yes | No | No |  |
| 9 | Kilifi | Yes | No | Female | 0-5 | Unknown | No | Resolved | No | No | No | No | No | No | No | Bronchiolitis |
| 10 | Kilifi | No | No | Male | 0-5 | Unknown | No | Resolved | No | No | No | No | No | No | No | Exacerbation of patent ductus arteriosus |
| 11 | Kilifi | No | No | Male | 0-5 | Unknown | Unlikely | Resolved | No | No | No | No | Yes | No | No | Suspected drug toxicity |
| 12 | Kilifi | No | No | Female | 36-40 | Unknown | No | Fatal | No | No | No | No | No | No | Yes |  |
| 13 | Kilifi | Yes | Yes | Male | 0-5 | Unknown | Unlikely | Fatal | No | No | No | No | Yes | No | No | Other encephalopathy |
| 14 | Kilifi | Yes | No | Female | 6-10 | Unknown | No | Resolved | No | Yes | No | No | No | No | No | Exacerbation of congenital heart disease |
| 15 | Kilifi | Yes | No | Female | 6-10 | Unknown | Unlikely | Resolved | No | Yes | No | No | No | No | No | Suspected drug toxicity |
| 16 | Kilifi | Yes | No | Female | 0-5 | Unknown | No | Resolved | No | Yes | No | No | No | No | No |  |
| 17 | Kilifi | No | Yes | Male | 11-15 | Negative | Unlikely | Resolved | No | No | Yes | No | No | No | No |  |
| 18 | Kilifi | Yes | No | Female | 0-5 | Unknown | No | Fatal | No | Yes | No | No | No | No | No |  |
| 19 | Kilifi | Yes | Yes | Female | 21-25 | Negative | Unlikely | Fatal | Yes | Yes | No | No | No | No | Yes | Epilepsy, aspiration |
| 20 | Kilifi | Yes | Yes | Female | 0-5 | Unknown | Unlikely | Resolved | No | Yes | No | No | No | No | No | Exacerbation of congenital heart disease |
| 21 | Kilifi | No | Yes | Female | 6-10 | Unknown | Unlikely | Resolved | No | No | No | Yes | No | No | No |  |
| 22 | Kilifi | Yes | No | Male | 6-10 | Negative | Unlikely | Fatal | No | No | No | Yes | No | No | No |  |
| 23 | Kilifi | Yes | Yes | Female | 6-10 | Unknown | Unlikely | Resolved | No | Yes | No | No | No | No | No | Exacerbation of congenital heart disease |
| 24 | Kilifi | No | Yes | Male | 0-5 | Unknown | Unlikely | Resolved | No | No | No | Yes | No | No | No | UTI |
| 25 | Kilifi | No | Yes | Male | 0-5 | Unknown | No | Resolved | No | No | No | No | No | No | No | UTI, Posterior Urethral Valve |
| 26 | Kilifi | No | Yes | Male | 6-10 | Unknown | Unlikely | Fatal | Yes | Yes | No | Yes | No | No | No | Other encephalopathy |
| 27 | Kilifi | Yes | No | Female | 0-5 | Unknown | Unlikely | Resolved | No | Yes | No | No | No | No | No |  |
| 28 | Kilifi | Yes | No | Female | 0-5 | Unknown | Unlikely | Fatal | Yes | Yes | No | No | No | No | No | HIV related |
| 29 | Kilifi | No | No | Male | 0-5 | Unknown | Unlikely | Resolved | No | Yes | No | No | No | No | No | Exacerbation of congenital heart disease |
| 30 | Kilifi | No | No | Male | 0-5 | Unknown | Unlikely | Fatal | No | Yes | No | No | Yes | No | No | Renal impairment |
| 31 | Kilifi | Yes | Yes | Female | 0-5 | Unknown | Unlikely | Fatal | No | Yes | No | No | No | No | No | HIV related |
| 32 | Nairobi | Yes | Yes | Male | 0-5 | Negative | Unlikely | Resolved | No | Yes | No | No | No | No | No |  |
| 33 | Nairobi | Yes | Yes | Male | 0-5 | Negative | No | Resolved | No | No | Yes | No | No | No | No |  |
| 34 | Nairobi | No | Yes | Female | 6-10 | Negative | No | Resolved | Yes | No | Yes | No | No | No | No | Exacerbation of congenital heart disease |
| 35 | Nairobi | Yes | No | Male | 11-15 | Negative | No | Fatal | No | No | No | No | No | No | No | Aspiration |
| 36 | Nairobi | Yes | No | Female | 20-25 | Negative | No | Fatal | Yes | No | No | Yes | Yes | No | No | Renal impairment |
| 37 | Nairobi | Yes | No | Female | 11-15 | Negative | Unlikely | Resolved | No | Yes | No | No | No | No | No | Aspiration |
| 38 | Nairobi | Yes | No | Female | 11-15 | Negative | Unlikely | Resolved | No | No | Yes | No | No | No | No |  |
| 39 | Nairobi | Yes | Yes | Female | 11-15 | Negative | Unlikely | Fatal | No | No | No | Yes | Yes | No | No | Probable meningitis |
| 40 | Nairobi | No | No | Male | 6-10 | Negative | Unlikely | Fatal | No | Yes | No | Yes | Yes | No | No |  |
| 41 | Kilifi | Yes | No | Female | 16-20 | Unknown | Unlikely | Resolved with sequelae | No | No | No | No | No | No | No | LP confirmed meningitis |
| 42 | Kilifi | No | No | Male | 41-45 | Unknown | Unlikely | Fatal | Yes | No | No | No | No | No | Yes | Epilepsy, soft tissue infection |
| 43 | Kilifi | No | No | Female | 46-50 | Unknown | Unlikely | Fatal | Yes | No | No | Yes | Yes | No | No | HIV related, Septic genital ulcers |
| 44 | Kilifi | Yes | No | Male | 20-25 | Negative | Unlikely | Resolved | No | No | No | No | No | No | No | Failed appetite test, severe malnutrition |
| 45 | Kilifi | Yes | Yes | Female | 16-20 | Infected | Unlikely | Fatal | No | No | No | No | No | No | No | Febrile illness unspecified, HIV exposed |
| 46 | Nairobi | No | Yes | Male | 51-55 | Negative | Unlikely | Fatal | Yes | Yes | No | No | Yes | No | Yes |  |
| 47 | Kampala | Yes | No | Female | 16-20 | Infected | Unlikely | Fatal | No | No | No | Yes | Yes | No | No | HIV related |
| 48 | Kampala | No | No | Female | 11-15 | Infected | Unlikely | Resolved | No | No | No | Yes | No | No | No |  |
| 49 | Kampala | No | No | Female | 11-15 | Infected | Unlikely | Resolved | No | No | No | Yes | No | Yes | No |  |
| 50 | Kampala | Yes | No | Male | 16-20 | Negative | Unlikely | Resolved | Yes | No | No | No | Yes | No | No | Febrile convulsions |
| 51 | Kampala | No | Yes | Female | 6-10 | Negative | Unlikely | Fatal | No | Yes | Yes | No | No | No | No |  |
| 52 | Kampala | Yes | No | Female | 11-15 | Negative | Unlikely | Resolved | Yes | No | No | No | Yes | No | No |  |
| 53 | Kampala | Yes | Yes | Female | 0-5 | Infected | Unlikely | Resolved | No | Yes | Yes | No | No | No | No | Exacerbation of congenital heart disease |
| 54 | Kampala | No | Yes | Female | 11-15 | Negative | Unlikely | Fatal | No | Yes | Yes | No | Yes | No | No |  |
| 55 | Kampala | Yes | Yes | Female | 6-10 | Negative | Unlikely | fatal | No | No | No | No | No | No | No | Febrile convulsions, probable meningitis |
| 56 | Kampala | Yes | Yes | Female | 36-40 | Negative | Unlikely | Fatal | No | Yes | No | No | Yes | No | Yes |  |
| 57 | Kampala | No | Yes | Male | 11-15 | Negative | Unlikely | Fatal | No | Yes | Yes | Yes | No | No | No |  |
| 58 | Kampala | Yes | Yes | Male | 11-15 | Infected | Unlikely | Fatal | No | No | Yes | Yes | Yes | No | No |  |
| 59 | Kampala | No | Yes | Female | 6-10 | Infected | Unlikely | Fatal | No | No | No | Yes | No | No | No |  |
| 60 | Kampala | Yes | No | Female | 11-15 | Negative | Unlikely | Resolved | No | Yes | No | Yes | No | No | No |  |
| 61 | Kampala | No | No | Female | 0-5 | Negative | Unlikely | Fatal | No | Yes | No | Yes | No | No | No |  |
| 62 | Kampala | Yes | Yes | Female | 6-10 | Negative | Unlikely | Fatal | No | No | No | Yes | No | Yes | No |  |
| 63 | Kampala | Yes | Yes | Female | 11-15 | Negative | Unlikely | Fatal | No | Yes | No | No | No | No | No |  |
| 64 | Kampala | No | Yes | Female | 11-15 | Negative | Unlikely | Fatal | No | Yes | No | No | Yes | No | No |  |
| 65 | Kampala | No | No | Male | 0-5 | Negative | No | Resolved | No | No | No | No | Yes | No | No | URTI |
| 66 | Kampala | Yes | Yes | Female | 11-15 | Negative | Unlikely | Fatal | No | No | No | Yes | No | Yes | No |  |
| 67 | Kampala | No | No | Female | 21-25 | Infected | Unlikely | Fatal | No | Yes | No | No | No | No | No |  |
| 68 | Kampala | Yes | No | Male | 11-15 | Negative | Unlikely | Fatal | Yes | No | No | Yes | Yes | No | No | Sickle cell disease |
| 69 | Kampala | No | Yes | Male | 26-30 | Negative | Unlikely | Resolved | No | No | No | No | No | No | No | Myelodysplastic disorder of down's syndrome |
| 70 | Kampala | Yes | Yes | Male | 31-35 | Negative | Unlikely | Fatal | No | Yes | No | No | Yes | Yes | No |  |
| 71 | Kampala | No | Yes | Male | 6-10 | Negative | Unlikely | Resolved | No | Yes | No | No | No | Yes | No | Exacerbation of congenital heart disease |
| 72 | Kampala | No | Yes | Male | 6-10 | Negative | No | Resolved | No | No | No | No | No | No | No | Exacerbation of congenital heart disease |
| 73 | Kampala | Yes | Yes | Male | 26-30 | Infected | Unlikely | Fatal | No | No | No | Yes | Yes | No | No |  |
| 74 | Blantyre | Yes | Yes | Male | 6-10 | Negative | No | Resolved | No | No | No | No | No | No | No | Epilepsy |
| 75 | Blantyre | Yes | Yes | Male | 6-10 | Negative | Unlikely | Resolved | No | No | No | No | No | No | No | Hydrocephalus |
| 76 | Blantyre | Yes | No | Male | 11-15 | Negative | No | Fatal | No | No | No | Yes | Yes | No | No |  |
| 77 | Blantyre | No | No | Male | 6-10 | Negative | No | Fatal | No | No | No | No | No | No | Yes | Aspiration |
| 78 | Blantyre | No | No | Male | 6-10 | Negative | Unlikely | Resolved | No | No | No | No | No | No | Yes | Laryngomalacia |
| 79 | Blantyre | No | Yes | Female | 11-15 | Negative | Unlikely | Resolved | No | No | No | No | Yes | No | No |  |
| 80 | Blantyre | No | No | Male | 0-5 | Negative | No | Fatal | No | No | No | Yes | Yes | No | No |  |
| 81 | Blantyre | Yes | No | Female | 6-10 | Negative | No | Fatal | No | No | No | No | No | No | No | Aspiration |
| 82 | Blantyre | Yes | No | Female | 0-5 | Negative | Unlikely | Resolved | No | Yes | No | Yes | Yes | No | No | Bronchiolitis |
| 83 | Blantyre | No | Yes | Male | 11-15 | Negative | No | Fatal | No | No | No | Yes | Yes | No | No |  |
| 84 | Blantyre | No | Yes | Male | 11-15 | Negative | No | Resolved | No | No | No | Yes | No | No | No |  |
| 85 | Blantyre | Yes | No | Female | 0-5 | Infected | Unlikely | Fatal | Yes | No | No | No | Yes | No | No |  |
| 86 | Blantyre | No | Yes | Male | 6-10 | Negative | Unlikely | Fatal | No | No | No | Yes | Yes | No | No |  |
| 87 | Blantyre | No | No | Male | 11-15 | Negative | Unlikely | Resolved | No | No | No | Yes | No | No | No |  |
| 88 | Blantyre | Yes | No | Male | 16-20 | Negative | No | Resolved | No | No | No | No | Yes | No | No |  |
| 89 | Blantyre | Yes | Yes | Female | 16-20 | Negative | No | Resolved | No | No | No | No | No | No | No | Bronchiolitis |
| 90 | Blantyre | No | Yes | Female | 26-30 | Negative | No | Resolved | No | No | Yes | No | No | No | No |  |
| 91 | Blantyre | No | No | Male | 11-15 | Negative | Unlikely | Fatal | Yes | No | No | No | Yes | No | No |  |
| 92 | Blantyre | Yes | No | Female | 11-15 | Infected | No | Resolved | No | No | Yes | No | No | No | No |  |
| 93 | Blantyre | Yes | Yes | Male | 6-10 | Infected | Unlikely | Fatal | No | Yes | No | Yes | No | No | No | HIV related |
| 94 | Blantyre | No | No | Female | 0-5 | Negative | No | Fatal | No | Yes | No | No | No | No | No |  |
| 95 | Blantyre | No | Yes | Female | 6-10 | Negative | No | Resolved | No | No | No | No | Yes | No | No |  |
| 96 | Blantyre | No | Yes | Female | 6-10 | Negative | No | Resolved | No | No | No | Yes | No | No | No |  |
| 97 | Blantyre | Yes | Yes | Female | 26-30 | Negative | No | Resolved | Yes | No | No | Yes | Yes | No | No |  |
| 98 | Blantyre | Yes | Yes | Female | 31-35 | Negative | No | Fatal | No | No | No | Yes | Yes | No | No |  |
| 99 | Blantyre | No | Yes | Female | 16-20 | Negative | No | Resolved | Yes | Yes | No | No | No | No | No |  |
| 100 | Blantyre | No | Yes | Male | 0-5 | Negative | No | Fatal | No | No | No | No | No | No | No | Community death |
| 101 | Blantyre | No | No | Male | 36-40 | Infected | Unlikely | Resolved | No | No | Yes | No | Yes | No | No | HIV related |
| 102 | Blantyre | Yes | No | Female | 16-20 | Infected | No | Fatal | No | No | No | No | No | No | No | HIV related, Hydrocephalus, Probable meningitis |
| 103 | Blantyre | Yes | Yes | Female | 11-15 | Negative | No | Fatal | No | No | No | No | Yes | No | No |  |
| 104 | Blantyre | No | No | Female | 31-35 | Infected | No | Fatal | Yes | No | Yes | No | Yes | No | No |  |
| 105 | Blantyre | No | Yes | Male | 11-15 | Negative | Unlikely | Fatal | No | Yes | No | Yes | No | No | No |  |
| 106 | Blantyre | Yes | No | Male | 26-30 | Infected | Unlikely | Fatal | No | No | No | No | Yes | No | No |  |
| 107 | Blantyre | No | No | Male | 11-15 | Negative | Unlikely | Fatal | No | No | No | No | Yes | No | No |  |
| 108 | Blantyre | No | Yes | Female | 21-25 | Negative | Unlikely | Fatal | No | No | No | Yes | No | No | Yes | Aspiration |
| 109 | Blantyre | No | Yes | Male | 26-30 | Negative | No | Resolved | Yes | No | No | Yes | No | No | No |  |
| 110 | Blantyre | Yes | Yes | Male | 21-25 | Negative | Unlikely | Fatal | No | No | No | Yes | Yes | No | No | Aspiration |
| 111 | Blantyre | Yes | Yes | Male | 21-25 | Negative | Unlikely | Resolved | Yes | Yes | Yes | No | Yes | No | No |  |
| 112 | Blantyre | Yes | Yes | Male | 21-25 | Negative | Unlikely | Resolved | No | No | No | No | Yes | No | No | Renal implant |
| 113 | Blantyre | Yes | Yes | Male | 21-25 | Negative | Unlikely | Resolved | No | Yes | Yes | No | No | No | No | Renal implant |
| 114 | Blantyre | Yes | Yes | Male | 51-55 | Negative | Unlikely | Fatal | No | No | No | Yes | Yes | No | No |  |
| 115 | Blantyre | No | Yes | Male | 26-30 | Negative | No | Resolved | No | No | No | No | Yes | No | No | Oral thrush |
| 116 | Blantyre | No | Yes | Male | 21-25 | Infected | No | Resolved | Yes | No | No | No | No | No | No |  |
| 117 | Blantyre | Yes | No | Male | 41-45 | Negative | Unlikely | Fatal | No | Yes | No | No | Yes | No | No |  |
| 118 | Dhaka | No | No | Female | 0-5 | Negative | No | Resolved | No | Yes | Yes | Yes | Yes | No | No |  |
| 119 | Dhaka | No | Yes | Female | 0-5 | Negative | No | Resolved | No | No | No | Yes | Yes | No | No |  |
| 120 | Dhaka | Yes | Yes | Male | 0-5 | Negative | No | Resolved | No | No | No | Yes | Yes | No | No |  |
| 121 | Dhaka | No | No | Female | 6-10 | Negative | No | Resolved | No | No | No | Yes | No | No | No | Severe acute malnutrition |
| 122 | Dhaka | No | No | Male | 0-5 | Negative | No | Resolved | No | No | No | Yes | No | No | No | Severe acute malnutrition |
| 123 | Dhaka | Yes | Yes | Female | 0-5 | Negative | No | Resolved | No | Yes | No | No | No | No | No | Severe underweight |
| 124 | Dhaka | No | No | Male | 0-5 | Negative | No | Fatal | Yes | Yes | No | No | No | No | No | Multisystem inflammatory syndrome, COVID-19 disease, heart failure, severe acute malnutrition |
| 125 | Dhaka | Yes | Yes | Male | 0-5 | Negative | No | Fatal | No | No | No | No | No | No | No | Sudden infant death syndrome, severe acute malnutrition |
| 126 | Dhaka | Yes | No | Male | 0-5 | Negative | No | Fatal | No | Yes | No | Yes | No | No | No | Inotropes refractory septic shock, severe acute malnutrition |
| 127 | Dhaka | Yes | No | Female | 0-5 | Negative | No | Resolved | No | No | No | No | No | No | No | Febrile illness unspecified |
| 128 | Dhaka | Yes | No | Female | 0-5 | Negative | No | Resolved | No | No | Yes | No | No | No | No |  |
| 129 | Dhaka | No | Yes | Female | 0-5 | Negative | Unlikely | Fatal | No | Yes | No | No | No | No | No | Ileus, Liver dysfunction, Inotropes refractory septic shock, hepatic encephalopathy, severe acute malnutrition |
| 130 | Dhaka | No | Yes | Male | 0-5 | Negative | No | Fatal | No | Yes | No | Yes | Yes | No | No | Ileus, Liver dysfunction, Inotropes refractory septic shock, severe acute malnutrition |
| 131 | Dhaka | No | Yes | Female | 0-5 | Negative | No | Resolved | No | No | Yes | No | No | No | No |  |
| 132 | Dhaka | No | Yes | Female | 0-5 | Negative | No | Resolved | No | Yes | Yes | No | Yes | No | No |  |
| 133 | Dhaka | Yes | Yes | Female | 0-5 | Negative | No | Resolved | No | Yes | No | No | No | No | No | Bronchiolitis |
| 134 | Dhaka | Yes | No | Male | 0-5 | Negative | No | Resolved | No | No | No | Yes | No | No | No |  |
| 135 | Dhaka | No | Yes | Male | 0-5 | Negative | No | Resolved | No | No | No | Yes | No | No | No | Late onset hospital acquired infection, severe acute malnutrition |
| 136 | Dhaka | Yes | No | Female | 0-5 | Negative | No | Fatal | No | No | No | Yes | Yes | No | No | Renal implant, Liver dysfunction, septic shock, sclerema, severe acute malnutrition |
| 137 | Dhaka | Yes | No | Female | 0-5 | Negative | No | Resolved | No | No | No | Yes | No | No | No | Hypocalcemia, urinary tract infection, severe acute malnutrition |
| 138 | Dhaka | Yes | Yes | Female | 6-10 | Negative | No | Resolved | No | No | Yes | No | No | No | No | Severe acute malnutrition |
| 139 | Dhaka | Yes | No | Female | 0-5 | Negative | No | Resolved | No | No | No | Yes | No | No | No |  |
| 140 | Dhaka | Yes | Yes | Female | 0-5 | Negative | No | Resolved | No | Yes | No | Yes | No | No | No |  |
| 141 | Dhaka | No | No | Male | 11-15 | Negative | No | Fatal | No | Yes | No | Yes | Yes | No | No | Multisystem inflammatory syndrome, disseminated intravascular coagulation, severe acute malnutrition |
| 142 | Dhaka | No | Yes | Male | 11-15 | Infected | No | Fatal | No | Yes | No | No | No | No | No | HIV related |
| 143 | Dhaka | Yes | Yes | Female | 0-5 | Negative | No | Resolved | No | No | No | Yes | No | No | No |  |
| 144 | Dhaka | No | Yes | Female | 0-5 | Negative | No | Fatal | No | Yes | No | No | Yes | No | No | Ileus, severe acute malnutrition |
| 145 | Dhaka | No | No | Female | 0-5 | Negative | No | Fatal | No | No | No | Yes | No | No | No | Severe acute malnutrition |
| 146 | Dhaka | No | No | Female | 0-5 | Negative | No | Resolved | No | Yes | No | Yes | Yes | No | No | Severe acute malnutrition |
| 147 | Dhaka | Yes | No | Male | 0-5 | Negative | No | Resolved | No | Yes | No | Yes | No | No | No | Moderate acute malnutrition |
| 148 | Dhaka | No | No | Male | 11-15 | Negative | No | Resolved | No | Yes | No | Yes | No | No | No | Severe acute malnutrition |
| 149 | Dhaka | Yes | Yes | Female | 0-5 | Negative | No | Fatal | No | Yes | No | Yes | Yes | No | No | Severe acute malnutrition |
| 150 | Dhaka | No | Yes | Male | 0-5 | Negative | No | Resolved | No | Yes | No | No | No | No | No | Severe acute malnutrition |
| 151 | Dhaka | No | No | Male | 6-10 | Negative | No | Resolved | No | Yes | No | No | No | No | No | Severe acute malnutrition |
| 152 | Dhaka | No | No | Male | 0-5 | Negative | No | Resolved | No | Yes | No | Yes | No | No | No | Severe acute malnutrition |
| 153 | Dhaka | No | No | Female | 11-15 | Negative | No | Fatal | Yes | Yes | No | Yes | Yes | No | No | Renal implant, ileus, Liver dysfunction, hypernatremia, severe acute malnutrition |

UTI; urinary tract infection, URTI; upper respiratory tract infection

**S7 Table.** Grade 3 & 4 toxicity events stratified by intervention groups.

| Type of toxicity | Pancreatic enzymes randomization |  | Bile acid randomization |  |
| --- | --- | --- | --- | --- |
|  | Placebo (n=216) | Pancreatic enzymes (n=213) | Placebo (n=217) | Bile acid (n=212) |
| All suspected toxicities | 2 | 2 | 2 | 2 |
| Hypovolemic shock | 1 | 1 | 1 | 1 |
| Diarrhea: increase of $\geq 7$ stools per 24h | 0 | 1 | 1 | 0 |
| Abnormal liver function (ALT $>10.0 \times$ ULN)* | 1 | 0 | 0 | 1 |

\*This child stopped trial interventions 16 days after enrolment.

**S8 Table.**
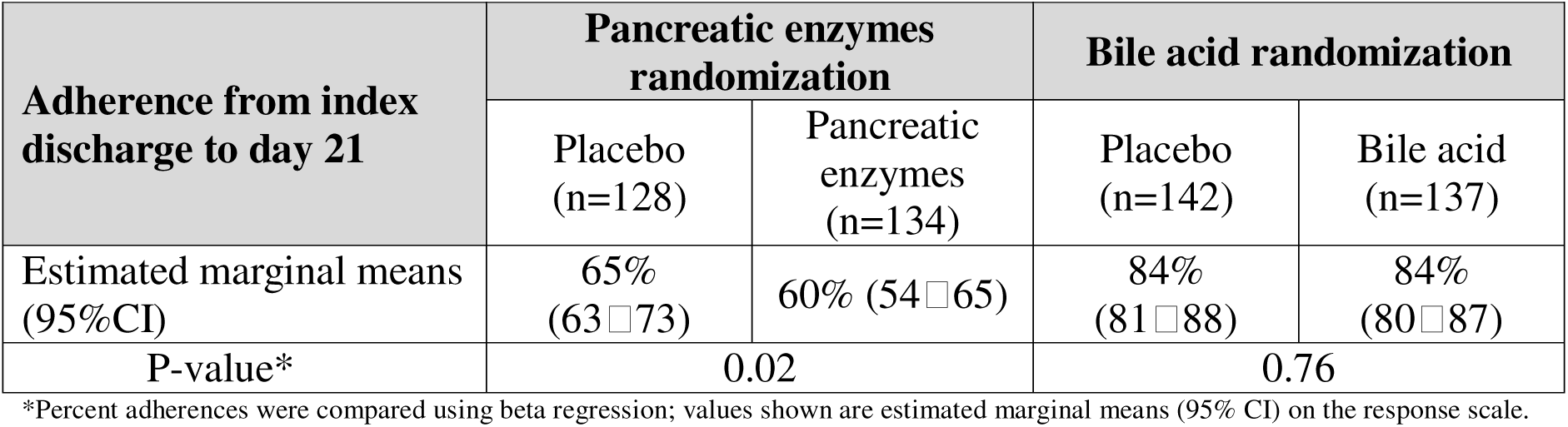
Adherence to study intervention after inpatient treatment stratified by intervention groups.

| Adherence from index discharge to day 21 | Pancreatic enzymes randomization |  | Bile acid randomization |  |
| --- | --- | --- | --- | --- |
|  | Placebo (n=128) | Pancreatic enzymes (n=134) | Placebo (n=142) | Bile acid (n=137) |
| Estimated marginal means (95%CI) | 65% (63□73) | 60% (54□65) | 84% (81□88) | 84% (80□87) |
| P-value* | 0.02 |  | 0.76 |  |

\*Percent adherences were compared using beta regression; values shown are estimated marginal means (95% CI) on the response scale.

**S1 Fig.**
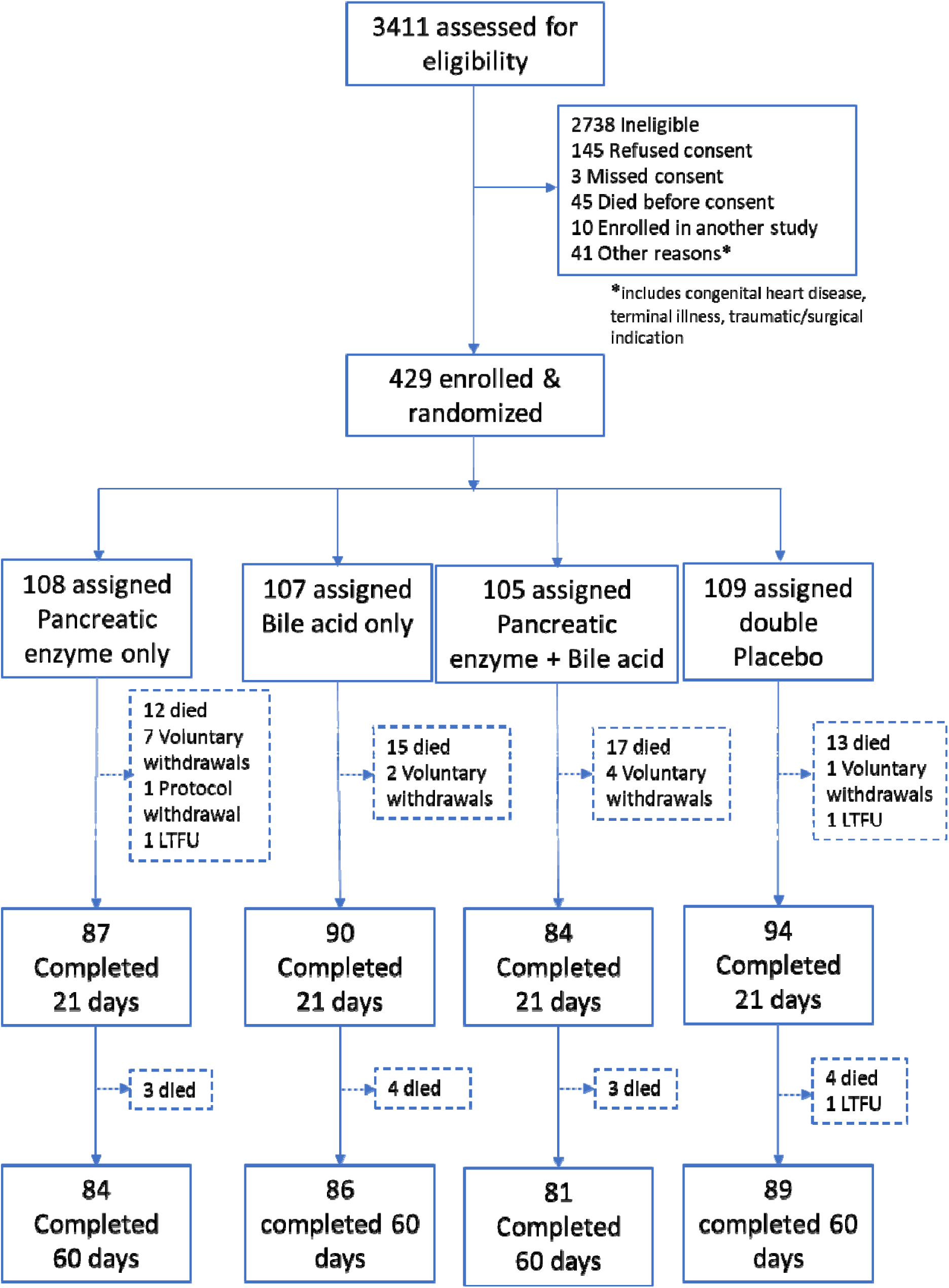
Trial participant flow chart stratified by four intervention groups.

**S2 Fig.**
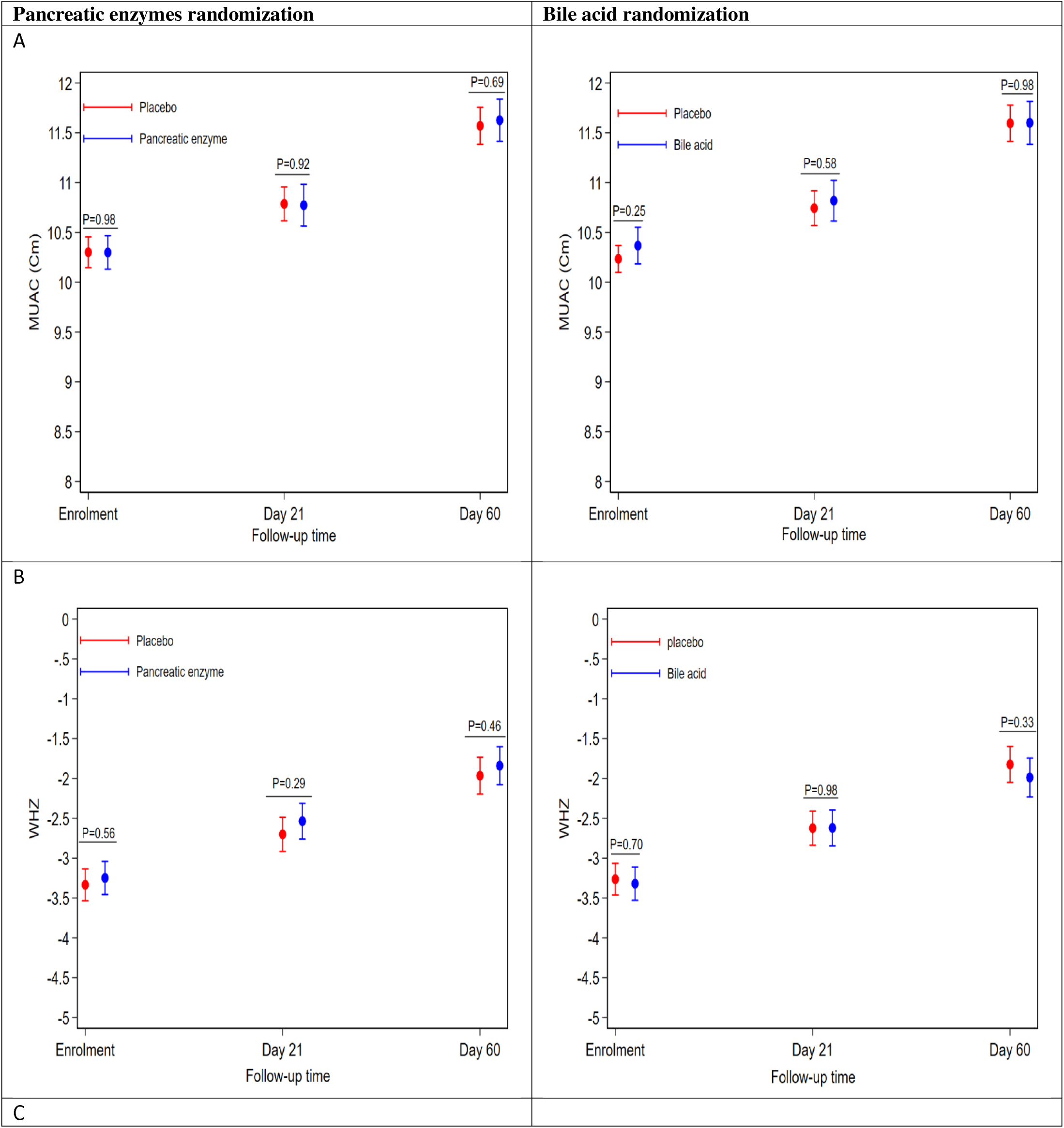
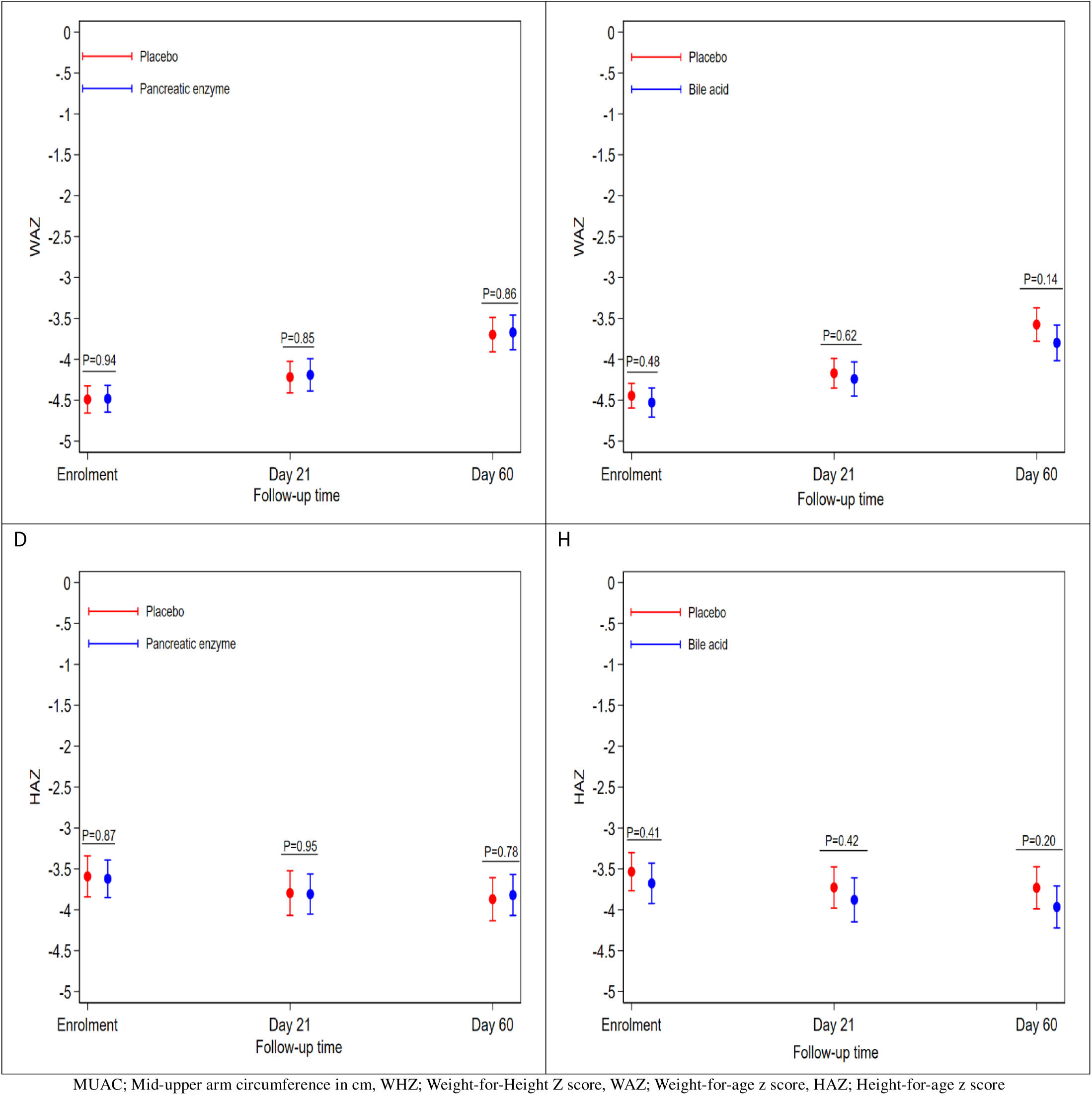
Absolute anthropometry at enrolment and follow-up by interventions: A. MUAC (Cm), B. WHZ, C. WAZ and D. HAZ.

## Glossary of terms and abbreviations

DSMB: Data & Safety Monitoring Board
IP: Investigational product(s)
KEMRI: Kenya Medical Research Institute
LMIC: Low- and Middle-Income Countries
MUAC: Mid-upper arm circumference
OxTREC: Oxford Tropical Research Ethics Committee
PI: Principal Investigator
RCT: Randomized controlled trial.
SAE: Serious adverse event
SAM: Severe acute malnutrition
SERU: KEMRI Scientific and Ethics Review Unit, Nairobi
SOP: Standard of operating procedure
SUSAR: Serious Unexpected Suspected Adverse Reaction
WHO: World Health Organization

## Introduction and background

Acutely ill children admitted to hospitals with severe malnutrition have high mortality and readmission rates, usually because of infection. Malnourished children have more potentially harmful bacteria in their upper intestines than well-nourished children and this may contribute to inflammation in the gut and whole body. These bacteria may cross from the intestines to the bloodstream causing life-threatening infections. A related abnormality among malnourished children is reduction in the digestive enzymes made by the pancreas and the liver. Apart from helping with digestion of food, these enzymes are important in helping the body control bacteria in the upper intestines. It is therefore possible that treatment with digestive enzymes could help reduce the burden of harmful bacteria and thus lower inflammation and the risk of serious infection. This trial aimed to find out if supplementing pancreatic enzymes and bile acids among ill children with severe acute malnutrition (SAM) is safe and reduces the risk of death, deterioration or readmission to hospital.

## Trial objectives

### Null hypothesis

Clinical outcomes amongst children admitted to hospital and treated for complicated SAM are not altered by administration of pancreatic enzymes or bile acids compared to placebo.

### Primary objective

To determine whether treatment with pancreatic enzymes or bile acids reduce mortality in acutely ill hospitalized children with SAM compared to placebo.

### Secondary objectives

To determine:

1. Rate and type of Severe Adverse Events (SAEs) (including readmissions to hospital).
2. Safety: rate of grade 3 or 4 toxicity events whilst receiving investigational products.
3. Intestinal function: number of days with diarrhoea during admission.
4. Use of second and third-line antibiotics during admission and readmission.
5. Number of days from enrolment to discharge during admission.
6. Growth: change in Mid-upper arm Circumference (MUAC), weight-for-length, weight-for-age and length-for-age z scores from enrolment to day 60.

## Trial design

### Overall Study Design

The trial investigated two separate non-antibiotic interventions in a 2×2 factorial, randomized, double-blind, controlled clinical trial (***Figure 1***). Two randomizations were performed. The two interventions will be analyzed separately. Evidence for interaction between the two interventions will be tested.

**Figure 1:**
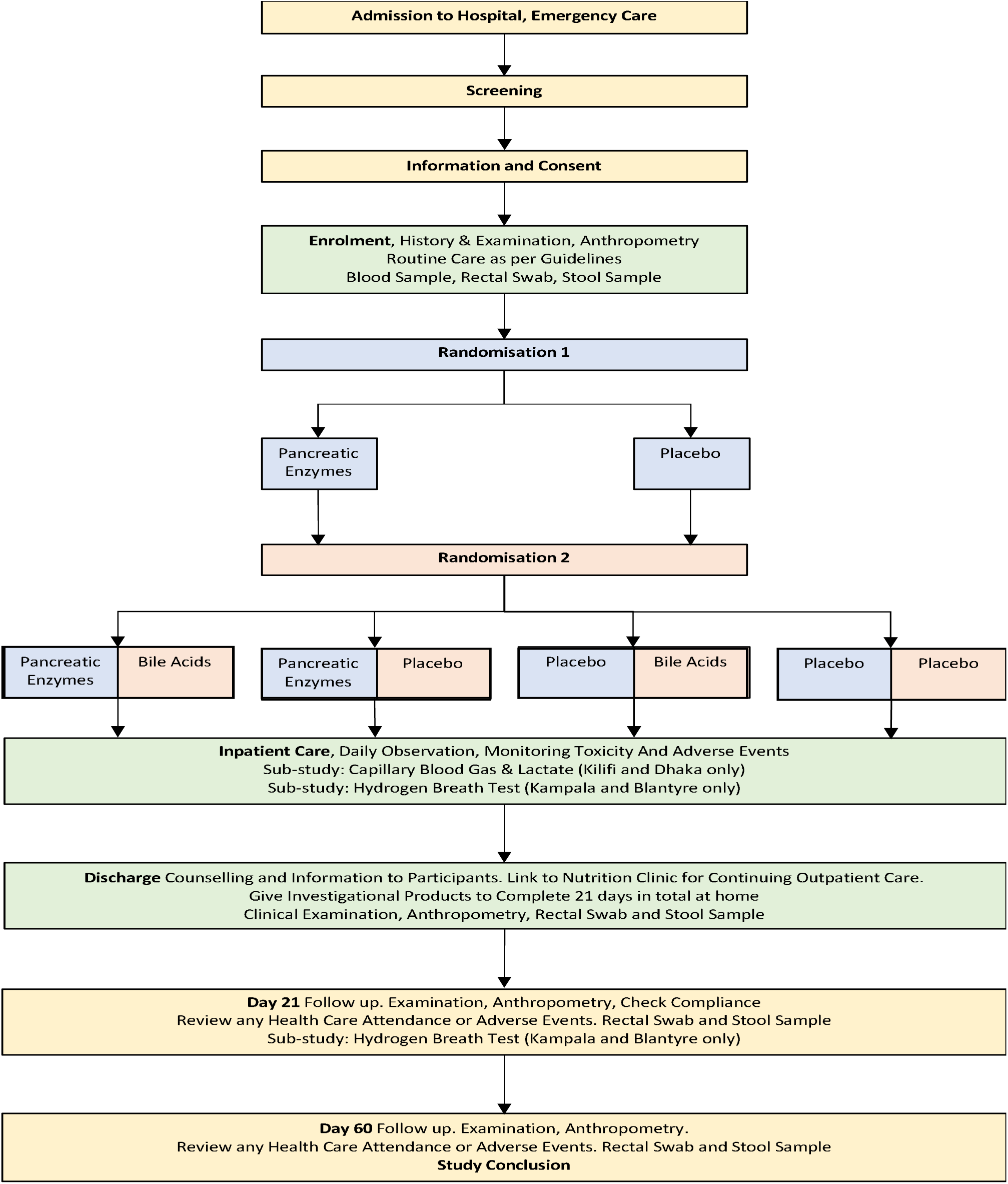
Trial scheme (Adopted from trial Protocol)

The trial was designed with three stages as below:

a. **Stage 1**, 200 participants were enrolled across all sites. Data Safety and Monitoring Board (DSMB) reviewed data for likelihood of major safety signals/harm (non-inferior to placebo) based on a composite endpoint of reported toxicity events, SAEs and deaths. A separate decision was made for each intervention. Both interventions were allowed to proceed to stage 2.
b. **Stage 2**, additional 200 participant were recruited giving a total of 400 participants. DSMB reviewed data on a composite endpoint of SAEs and deaths and results of modelled simulations for the likelihood of superiority over placebo for mortality. Interventions with less than 80% probability of efficacy were discontinued. A separate decision was made for each intervention arm. All interventions were discontinued due to lack of simulated efficacy.
c. **Stage 3**, a further 800 participants were to be enrolled with a total target sample size of 1,200 participants across all sites, based on a hazard ratio of 0.66 (a 33% reduction in mortality) from a baseline mortality of 24% to 16% with 90% power, 20% inflation for the factorial design and an estimated 5% loss to follow up.

### Recruitment & sites

Participating sites were: Kilifi County Hospital, Kenya; Coast General Hospital, Mombasa, Kenya; Mbagathi Sub-County Hospital, Nairobi, Kenya; Migori County Hospital and Ombo Mission Hospital, Migori, Kenya; Queen Elizabeth Central Hospital (QECH) Blantyre, Malawi; the International Centre for Diarrhoeal Disease Research Hospital, Dhaka, Bangladesh (ICCDR-B); and Mulago National Referral Hospital, Kampala, Uganda.

### Trial population

The trial population were children aged between 2 months to 59 months presenting to hospital with complicated severe acute malnutrition.

### Inclusion criteria

- Age ≥2 to < 60months
- Admitted to hospital with an acute, non-traumatic illness and within 72 hours of admission at the time of enrolment.
- Severe malnutrition (weight-for-height <-3 z scores of the median WHO growth standards and/or mid upper arm circumference <115mm (<110mm age below 6 months), or symmetrical oedema of at least the feet related to malnutrition (not related to a primary cardiac or renal disorder)
- Presence of two or more features of severity as specified in ***Table 1*** below. If a child met two criteria, they were enrolled before further criteria was assessed (e.g. a child was eligible on clinical signs before the complete blood count results are known).
- Accompanied by care provider who provided written informed consent.
- Primary caregiver plan to stay in the study area during the duration of the study.

**Table 1.**
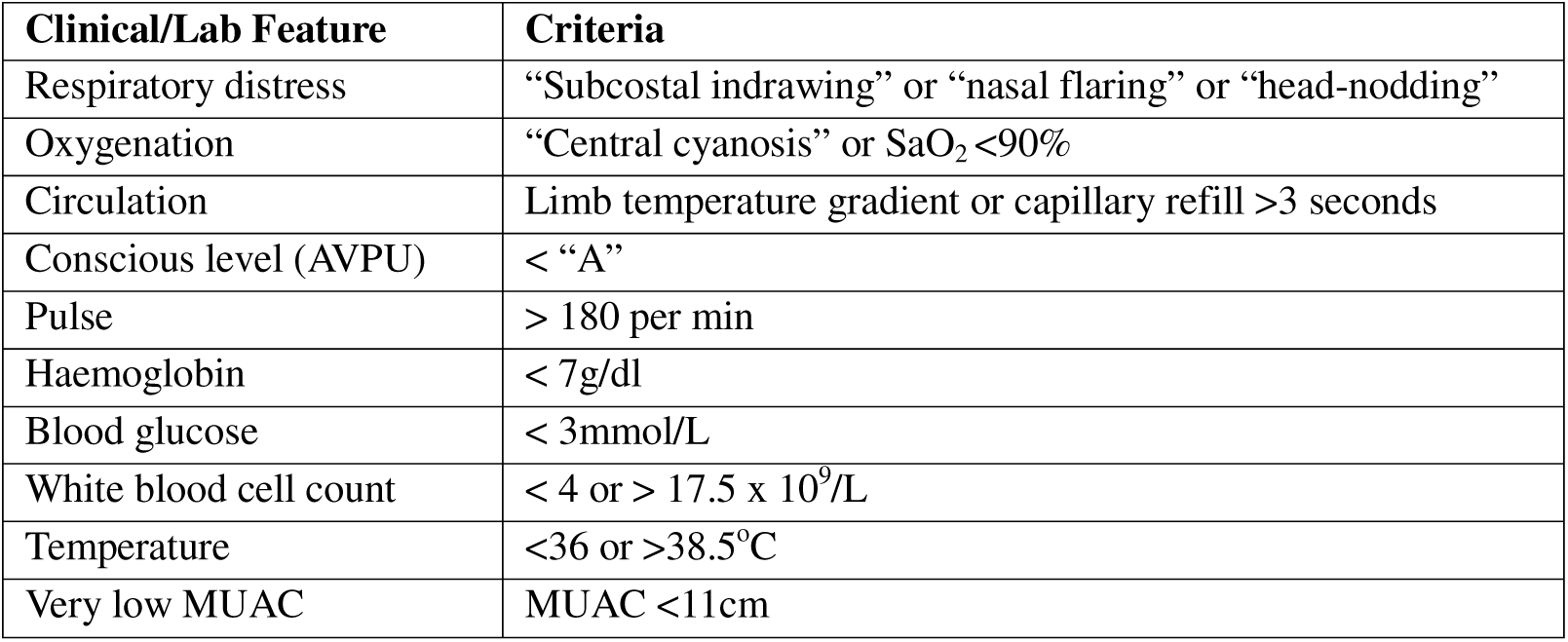
Severity features, two or more are required for enrolment. (Adopted from study Protocol)

### Exclusion criteria

- Requires immediate cardiac/respiratory resuscitation (may be re-evaluated later).
- Presence of terminal illness (other than severe acute malnutrition) likely to result in death within 6 months in the opinion of the recruitment team.
- Known congenital heart disease.
- Admission for traumatic or surgical indication.
- Known liver disorder or exocrine pancreatic disorder – e.g. biliary atresia, history of gallstones, cystic fibrosis or clinical jaundice.
- Known stomach or duodenal ulcer.
- Residence is outside the catchment area of each study hospital.
- Primary caregiver declines to provide informed consent.
- Known intolerance or allergy to any study medication.

## Statistical Methods

### Analysis population

The Intent-to-Treat (ITT) Population will be used in the analyses. The ITT Population is defined as all randomized subjects ignoring noncompliance, protocol deviations, withdrawal, and anything that happens after randomization.

### Estimation, Confidence Intervals and Hypothesis Testing

Estimates of treatment effects will be reported accompanied by 95% confidence intervals. Two-sided tests of statistical significance will be used. Significance level will be at P-value <0.05.

### Missing data

For demographic, clinical, social and laboratory variables, data will be assumed missing not at random and a “missing” category will be added, or in the case of HIV status, a “not tested” category.

For anthropometry, missing length/height values will be expected from participants who have chronic illness such as congenital abnormality or are too sick for their length/height measurement to be taken. Height/length at discharge may be used in place of admission values when accurate measurement at admission are missing. At follow up, missing anthropometric data are expected in participants who missed their visits or confirmed their vital status by phone only. The secondary outcome on growth, will only include children with anthropometry at day 21 or 60 of follow-up.

### Trial reporting

Reporting of trial results will follow the CONSORT guidelines for reporting outcomes in Trials^1^. Analysis of the two interventions will be conducted separately comparing each intervention to placebo. An analysis testing interaction between the two interventions will be conducted.

## Statistical Analysis

### Baseline Characteristics

Child demographic (age & sex), clinical and laboratory characteristics at enrolment will be reported by interventions separately in table 1. Categorical data will be summarized using frequency and proportions (%). Proportion of missing data will be reported. Continuous data will be reported as mean and standard deviation or median and Interquartile range (IQR) for variables with skewed distribution.

### Participant flow

Figure 1 will report participants screened, enrolled, randomized to each intervention, attrition from each group and those who completed day 60 of follow-up.

Child-years under observation from enrolment to day 60 or last date in the trial stratified by each intervention with 95% confidence intervals will be reported.

### Primary Outcome

The primary outcome is 60-day all-cause mortality. Absolute number of deaths in each arm, the mortality rate and 95% Confidence intervals will be reported.

The primary outcome efficacy will be assessed using difference in mortality rate (95% CI) and hazard ratio (95% CI) of death up to 60 days after enrolment calculated using Cox Proportion survival regression method and visualized using a Kaplan-Meier curve (Figure 2a & 2b). A sensitivity analysis will control for recruiting site to account for randomization being stratified by site. Appropriate survival regression models like shared frailty model or multilevel models (with site as a random effect) will be used.

Test of interaction including the two randomization arms as interaction term in the analysis of mortality will be performed using log-likelihood ratio test and reported.

Causes of death adjudicated by an endpoint review panel will be tabulated by allocated groups in supplemental material.

### Pre-specified Secondary Outcomes

The following secondary endpoints will be assessed:

- Incidence rates of SAEs will be estimated and compared between each intervention and placebo using incidence rate ratios.
- Episodes and incidence rates of suspected grade 3 and 4 toxicity events during 21 days from enrolment (when receiving study intervention) will be estimated and compared between each intervention and placebo using incidence rate ratios. Any toxicity events will be tabled as supplemental material.
- Incidence rates of readmission from discharge to day 60 will be estimated and compared between each intervention and placebo using incidence rate ratios.
- Days spent in hospital, days with signs of sepsis, diarrhoea and on antibiotics will be calculated, reported as median (IQR) and compared using appropriate count model (i.e Poisson, negative binomial or zero-inflated version if large proportion have zero counts).
- Changes in MUAC; weight-for-length; weight-for-age and length-for-age from baseline to 21 and 60 days after enrolment will be calculated and measure of effect estimated from a multivariable linear regression model with the difference as dependent variable and the randomization arm as independent variable adjusted for the absolute baseline values^2^.

## Statistical software

All statistical analysis will be conducted using use STATA version 17.0 (StataCorp, College Station, TX, USA) and R version 4.0.2.

